# Food-level dietary factors reveal cardiometabolic risk hidden by diet quality scores

**DOI:** 10.64898/2026.11.10.26367678

**Authors:** Hannah D. Kittrell, Justin Kauffman, Roelof A.J. Smit, Natalie N. Atabaki, Jordi Merino, Ryan W. Walker, Ruth J.F. Loos, Girish N. Nadkarni

## Abstract

**Introduction:** Diet quality scores are well-established predictors of cardiometabolic health but compress an individual’s food intake into a single dimension, potentially masking dietary variation relevant to disease risk. Whether this hidden variation carries prognostic information has not been tested at scale.

**Methods:** We applied non-negative matrix factorization (NMF) to the reported intake of 148 individual foods and beverages from 87,162 UK Biobank participants. We tested the association of NMF-derived intake factors with incident coronary artery disease (CAD), type 2 diabetes (T2D) and chronic kidney disease (CKD) over a median 9.4 years of follow-up using Cox proportional hazards models. We evaluated the prognostic value of the factors beyond diet quality scores using stepwise Cox models and net reclassification improvement (NRI), and characterized their metabolic profiles through metabolome-wide association analyses (MWAS) of 249 circulating metabolites.

**Results:** NMF identified eight reproducible intake factors that were stable across geographically distinct samples. Four factors were significantly associated with incident cardiometabolic disease: a high sugar and processed foods factor was adversely associated with T2D (HR 1.19; 95% CI 1.11-1.27) and CKD (HR 1.15; 1.07-1.23), while a plant-based and fish factor was protective for CKD (HR 0.90; 0.84-0.96) and a cheese and whole grains factor was protective for CAD (HR 0.94; 0.90-0.98). The factors improved disease discrimination beyond both diet quality scores (ΔC up to +0.0049) and correctly reclassified 8–16% more individuals, whereas adding either score to models containing the factors produced negligible further improvement. The high sugar and processed foods factor was associated with depletion of omega-3 fatty acids and the plant-based and fish factor showed enrichment in polyunsaturated fatty acids.

**Conclusions:** Food-level dietary intake profiling captures clinically relevant variation that established diet quality scores mask, offering a path toward more personalized dietary assessment for cardiometabolic disease prevention.

---

Cardiometabolic diseases, including coronary artery disease (CAD), type 2 diabetes (T2D) and chronic kidney disease (CKD), are the leading causes of morbidity and mortality worldwide, accounting for a combined 12.5 million deaths in 2023^1^. Suboptimal diet is a leading modifiable risk factor for these diseases,^2–4^ yet adherence to population-level dietary guidelines remains low^5–7^. One reason may be that while current guidelines offer some advice based on life stage and specific health conditions, they do not account for an individual’s behavior, biology or preferences^8^. Personalized dietary guidance may improve long-term adherence, but it requires methods that describe what an individual actually eats, including which foods are consumed together, rather than only how healthy their diet is overall^9,10^.

A central challenge in nutritional epidemiology is how best to assess and represent dietary intake. *A priori* diet quality scores like the Alternative Healthy Eating Index-2010^11^ (AHEI) and alternate Mediterranean Diet score^12^ (aMED) quantify and evaluate the healthfulness of a person’s diet by in a single number^11–13^. Because two people can reach the same score through different combinations of foods, a single score may conceal disease-relevant differences between them; whether this hidden variation carries prognostic information has not been tested at scale^14^. Data-driven methods, including principal component analysis (PCA), factor analysis and clustering^14,15^, have been developed to derive empirical dietary patterns from food intake data ^14^. However, these methods yield patterns with both positive and negative food weights that are hard to interpret, typically assign each individual to one dominant pattern and usually require foods to be collapsed into broad groups prior to analysis^14–16^.

Non-negative matrix factorization (NMF) is a representation learning method that identifies groups of foods that tend to be eaten together, directly from each person’s reported intake. As food intake is naturally non-negative (people eat zero servings or more of each food) and often sparse (many foods are eaten only occasionally), NMF suits the structure of dietary data^17,18^.

Importantly, NMF represents each person’s dietary intake as a blend of multiple intake factors rather than assigning them to a single dietary pattern, and it allows the contribution of individual foods to be traced through each factor, properties that may facilitate more personalized dietary guidance^9,18^. Previous applications of NMF in nutritional epidemiology identified dietary patterns and described their characteristics but did not test whether these patterns predicted health outcomes over time, reduced the food intake data into food groups before analysis and assigned individuals to a single pattern^22–24,46^.

To test whether food-level dietary patterns capture clinically relevant variation that diet quality scores miss, we applied NMF to 148 foods and beverages reported by 87,162 UK Biobank (UKB) participants. We derived eight intake factors in a discovery sample and confirmed their stability in held-out replication and geographically distinct validation samples. We related these factors to incident CAD, T2D and CKD over a median follow-up of 9.4 years, tested whether they added prognostic information beyond the AHEI and aMED, estimated their potential population impact and characterized their metabolic profiles. Finally, we compared individuals with near-identical diet quality scores but different factor profiles and traced differences in their predicted risk to specific foods.

## Results

This study included 87,162 UKB participants with at least two valid 24-hour dietary recalls (Fig. 1a). Participants had a mean age (SD) of 59.2 (7.8) years, mean body mass index (BMI) of 26.3 (4.3) kg/m^2^ and 57.0% were female (Extended Data Table 1). Over a median follow-up of 9.4 years, we observed 3,459 (4.0%) incident CAD events, 1,452 (1.7%) incident T2D events and 1,723 (2.0%) incident CKD events.

**Figure 1.**
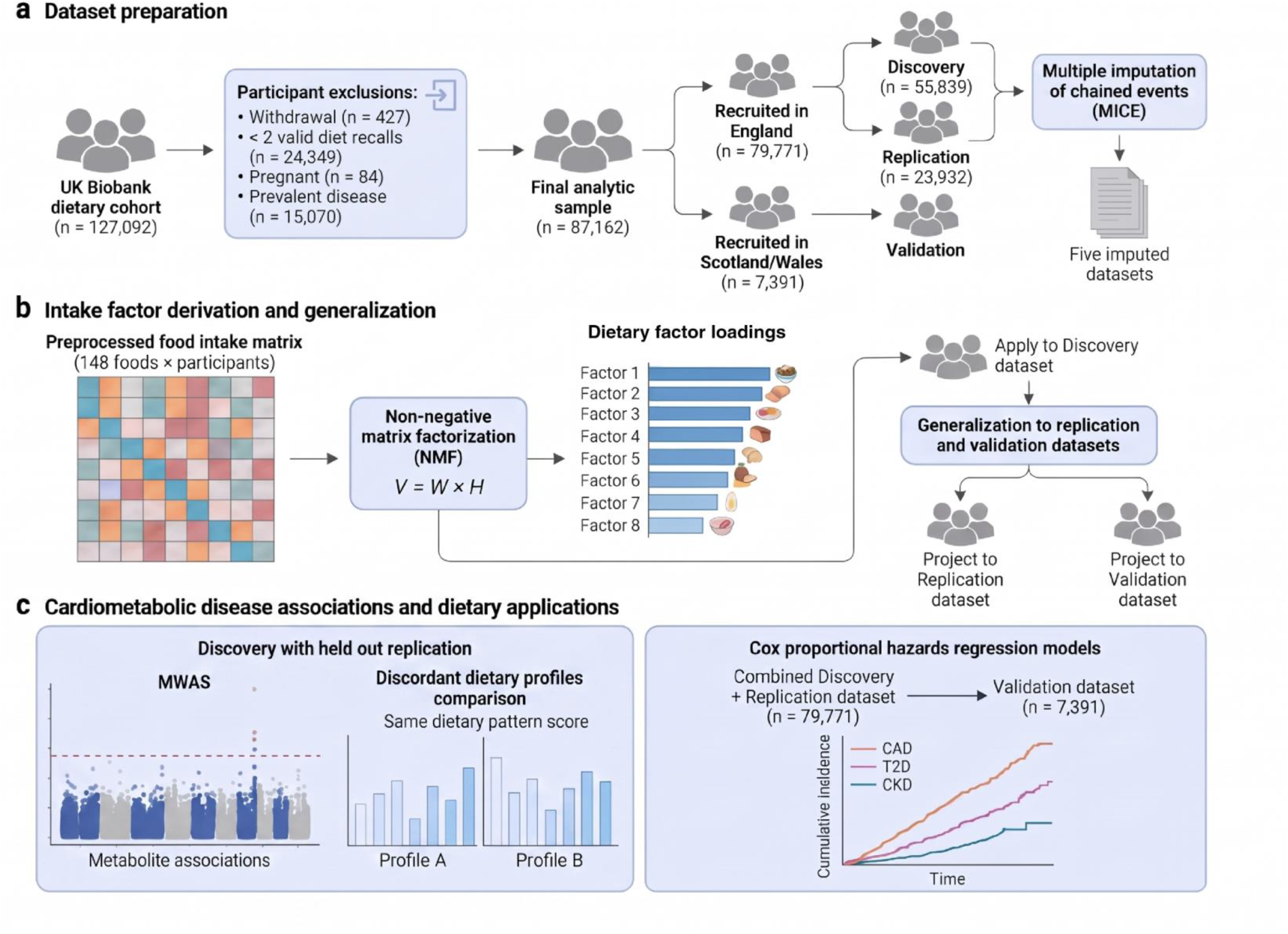
Study overview. **a,** Dataset preparation. From 127,092 UK Biobank participants with valid dietary intake data, 87,162 met inclusion criteria. After participant exclusions, UKB participants recruited in England were randomly split (70/30) into discovery and replication datasets and participants recruited in Scotland and Wales formed a geographically independent validation dataset. Missing covariate data were imputed separately within each group using multiple imputation by chained equations (MICE), generating five imputed datasets for each dataset. **b,** Intake factor derivation and generalization. NMF was applied to a preprocessed food intake matrix of 148 foods in the discovery dataset. Factor structure was assessed by de novo NMF replication in the replication and validation datasets, and individual factor loadings were obtained in held-out datasets by H-matrix projection. **c**, Downstream analyses. Left: metabolome-wide association study (MWAS) and discordant dietary profile analysis were conducted in the discovery dataset with testing in held-out replication dataset. Right: Cox proportional hazards regression models for incident CAD, T2D and CKD were fitted in combined England (*n* = 79,771) dataset using attained age as the timescale

### Eight intake factors replicate across independent samples

We applied NMF to the reported intake of 148 individual foods and beverages in a discovery dataset of 55,839 UKB participants recruited in England (70% random sample; Fig. 1b). The method identified groups of foods that are commonly consumed together, which we refer to as intake factors, without requiring foods to be aggregated into pre-defined groups^19^. A structured selection process determined that eight factors best balanced the ability to represent individual variation in dietary intake with the stability and interpretability of the factors (Methods; Supplementary Figs. 1-4). The remaining participants recruited in England formed a replication dataset (n = 23,932) and participants recruited in Scotland and Wales formed a geographically independent validation dataset (*n* = 7,391; Fig. 1a).

The eight factors spanned a wide range of dietary intake behaviors (Fig. 2a), with a mean of 11.3 active foods per factor and a median of 43% of foods that were non-zero but below the active threshold (Methods; Supplementary Table 9). **Factor 1 (F1; prudent base)** was the broadest, defined by tea, high-fiber fruits, whole grains and low-fat dairy. **Factor 2 (F2; cheese and whole grains)** emphasized coffee, hard and soft cheese and whole grains. Three factors were vegetable-focused: **Factor 3 (F3; meat and starchy vegetables)** combined starchy and root vegetables, leafy greens and red meat; **Factor 5 (F5; aromatic vegetables and rice)** captured cooking aromatics (garlic, onions, mushrooms) and simple starches such as white rice and pasta; **Factor 8 (F8; raw salad)** mapped onto salad greens and vegetables typically eaten uncooked. **Factor 4 (F4; high sugar and processed)** concentrated on refined grains, added sugar and sugar-sweetened beverages (SSBs). **Factor 6 (F6; fruit and LF dairy)** was defined by tropical fruits (grapes, berries, melons, pineapple) and low-fat yogurt. **Factor 7 (F7; plant-based and fish)** was a nutrient-dense profile defined by plant-based fats (nuts, seeds, avocado), spinach and oily fish. Notably, foods from single food groups (e.g., vegetables) were active across multiple factors, reflecting the diversity of culinary contexts in which individual foods are consumed. Inter-factor correlations were low (maximum |*r*| = 0.35), suggesting that the factors captured distinct dimensions of dietary intake (Fig. 2b).

**Figure 2.**
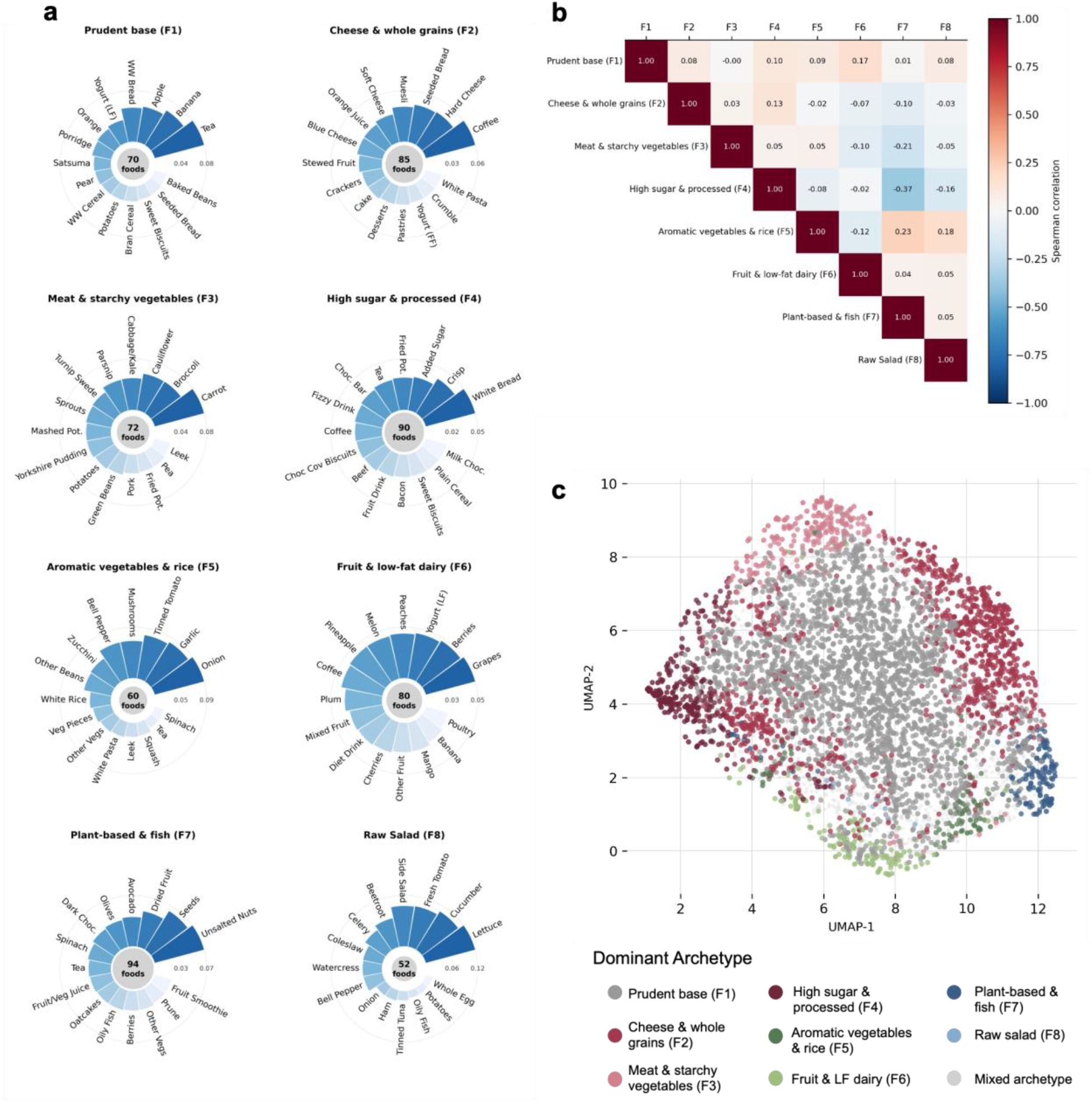
Food composition of eight latent intake factors, inter-factor correlations and archetype assignment. **(a)** Radial bar plots show the top-weighted foods for each of the eight NMF-derived intake factors. The top 15 foods are shown per factor, ranked by relative factor weight; bar length is proportional to the percent contribution to total factor weight. The central circle is sized proportionally to the number of non-zero foods assigned to each factor (range 42–87). Factor labels were assigned descriptively from each factor’s active foods. **(b)** Heatmap displaying pairwise Spearman correlation coefficients between food loadings (H-matrix) for the eight factors. The color scale indicates correlation strength, ranging from blue (negative) to dark red (positive). **(c)** 2D UMAP projection of NMF archetype assignment across the discovery dataset (n=55,839). Participants whose dominant factor accounted for more than 40% of their total factor loadings were assigned to that factor’s archetype; all others were classified as Mixed. Each point represents an individual, colored according to their NMF archetype. LF, low-fat.

To confirm that the factors were not specific to the discovery sample, we tested their reproducibility in two ways (Methods). First, we independently derived factors in the replication and validation datasets. All eight factors reproduced in the replication dataset, with high agreement in food composition (top-10 food overlap per factor of 67–100%; Extended Data Fig. 1). In the geographically independent validation dataset, six of eight factors reproduced; the prudent base (F1) and plant-based and fish (F7) factors did not, likely reflecting regional differences in food culture between England and Scotland/Wales. Second, we applied the discovery-derived factor structure to new participants and found that it explained dietary variation equally well in the replication dataset and with only a modest decrease in the validation dataset (Methods; Supplementary Table 10). Together, these results indicate that the eight factors capture reproducible patterns of dietary intake.

### Most participants’ diets combine several intake factors

NMF represents an individuals diet as a blend of all eight factors. To descriptively characterize this variation, we normalized factor loadings to proportional contributions and assigned individuals to archetypes based on their dominant factor loading (Methods). Most participants (52%) had a Mixed archetype, meaning no single factor accounted for a large share of their dietary intake (Fig. 2c; Supplementary Table 11). Among participants with a dominant factor, those in the plant-based and fish (F7) archetype were predominantly female (78.4%), had the highest diet quality scores and lowest BMI (24.4 kg/m^2^), whereas the high sugar and processed foods (F4) archetype was predominanly male (70.8%) had the lowest diet quality scores and highest BMI (28.1 kg/m^2^; Extended Data Table 1). Factor–diet quality score correlations were modest (maximum |*r*| = 0.37) and two factors, F3 (meat and starchy vegetables) and F8 (raw salad), showed little correlation with either score, suggesting they capture variation in dietary intake not well represented by the aggregate AHEI and aMED scores (Supplementary Fig. 5). The combination of an individual’s primary and secondary factor loading was also associated with their AHEI and aMED scores (Supplementary Fig. 6).

### Intake factors are associated with incident cardiometabolic diseases

We tested each factor in relation to incident CAD, T2D and CKD using Cox proportional hazards models adjusted for demographic, socioeconomic and lifestyle covariates in the combined sample of participants recruited in England (n = 79,771; Methods; Fig. 1c). Hazard ratios (HRs; pooled across imputed datasets) were expressed per 1-SD higher factor loading, with all eight factors entered simultaneously. Three factors had FDR-significant associations (Fig. 3a). The high sugar and processed foods factor (F4) had the strongest adverse associations, with HRs (95% CIs) of 1.19 (1.11–1.27; *P* = 9.2 × 10^−6^) for T2D and 1.15 (1.07–1.23; *P* = 5.2 × 10^−5^) for CKD. Individuals in the highest tertile of F4 loading consumed approximately 1.0 more serving/day of white bread, 0.6 more of added sugar, 0.3 more of crisps and 0.2 more of fried potatoes than individuals with zero loading on F4 (Supplementary File 6). The plant-based and fish factor (F7) showed the strongest protective association, with a HR of 0.90 (0.84–0.96; *P* = 8.9 × 10^−4^) for CKD. The highest-tertile F7 consumers ate approximately 0.25 more servings/day of unsalted nuts and 0.11 more of seeds than individuals with zero loading on F7 (Supplementary File 6). The cheese and whole grains factor (F2) was protective for CAD (HR 0.94, 0.90–0.98; *P* = 0.003). Several additional associations were nominally significant, including F5 (aromatic vegetables and rice) for T2D (HR 1.07, *P* = 0.015), F8 (raw salad) for T2D (HR 0.94, *P* = 0.041) and F2 for CKD (HR 0.94, *P* = 0.045).

**Figure 3.**
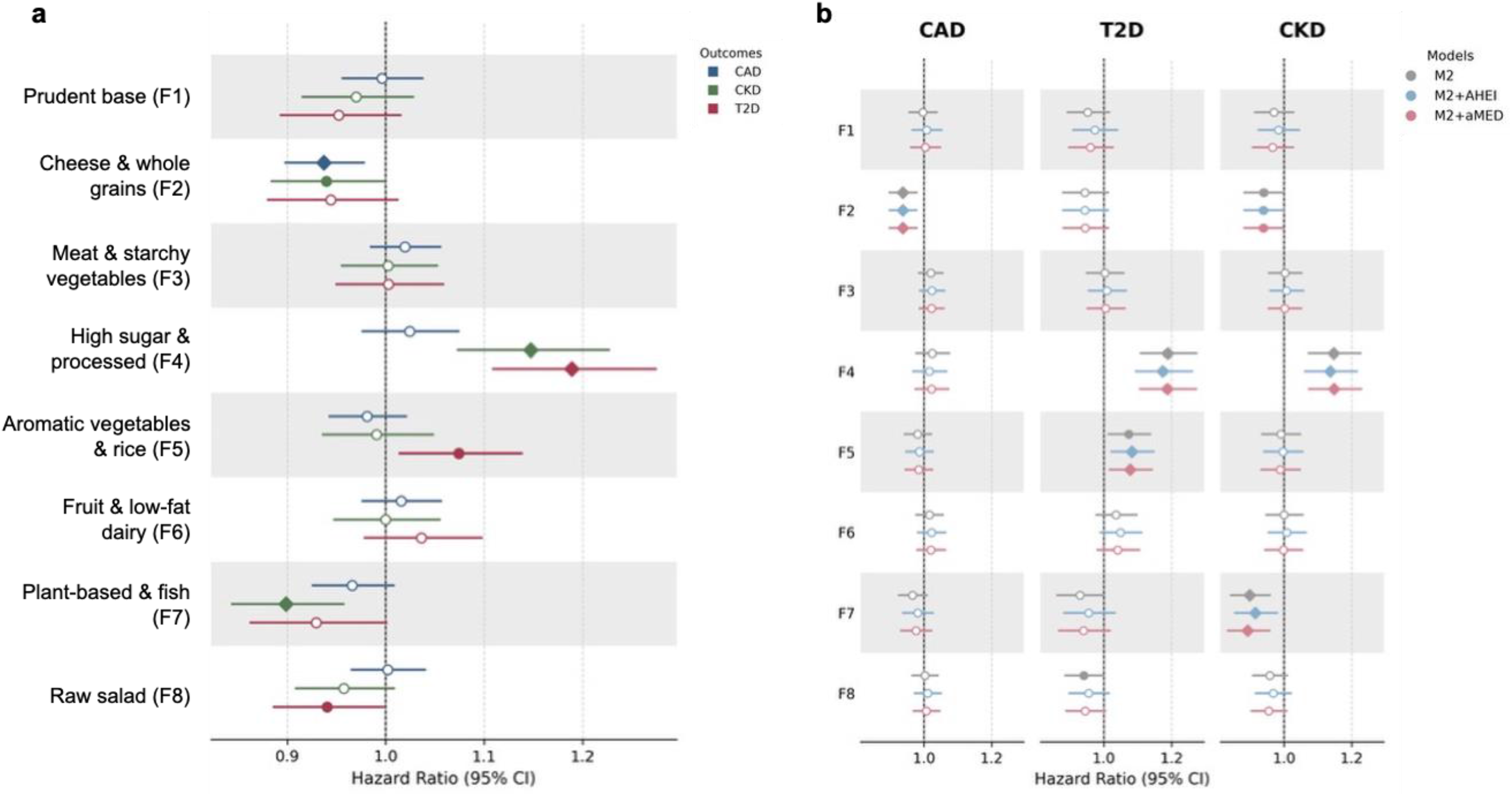
Associations of intake factors with incident cardiometabolic disease. **(a)** Forest plot showing hazard ratios (HRs) and 95% confidence intervals (CIs) per 1-SD increase in z-scored loading for each NMF-derived intake factor (F1-F8) in relation to coronary artery disease (CAD; blue), type 2 diabetes (T2D; red) and chronic kidney disease (CKD; green) in the combined discovery + replication datasets (*n* = 79,771). All eight factors were entered simultaneously into Cox proportional hazards models using attained age as the timescale and adjusted for Model 2 (M2) multivariable covariates (total energy intake, nation, ethnicity, education, income, smoking status, alcohol intake, physical activity, BMI, supplement use, family history of diabetes and family history of heart disease). Results were pooled across five multiply imputed datasets using Rubin’s rules. **(b)** Comparison of HRs before and after incremental adjustment for diet quality scores: the primary multivariable model (M2; gray), M2 additionally adjusted for the Alternative Healthy Eating Index-2010 (M2+AHEI; blue), and M2 additionally adjusted for the alternate Mediterranean Diet score (M2+aMED; red) across CAD, T2D and CKD. In both panels, filled diamonds denote FDR-significant associations (*P* <0.05 after Benjamini-Hochberg correction within each outcome), filled circles represent nominally significant associations and open circles represent non-significant associations.

Results were robust in pre-specified sensitivity analyses, including extended washout periods, restriction to participants with three or more dietary recalls, and compositional regression models that account for the substitution structure of dietary intake (Supplementary Files 11-12). E-values for FDR-significant associations ranged from 1.33 to 1.66, indicating that unmeasured confounding would need to be at least moderate to explain these findings (Supplementary Table 12). Pre-specified subgroup analyses tested interactions with sex, age, BMI, smoking status and ethnicity; no interaction survived correction for multiple testing (9 of 120 tests nominally significant at *P* < 0.05, none at Bonferroni-corrected *P* < 4.2 × 10^−4^; Supplementary Files 9-10).

Random survival forests (RSF) ranked the high sugar and processed foods factor (F4) as the most important dietary factor for T2D (7th of 23 predictors) and CKD (4th), and the meat and starchy vegetables factor (F3) as the most important dietary factor for CAD (10th; Methods; Supplementary File 14).

### Intake factors add prognostic value beyond established diet quality scores

To test whether the factor–disease associations were independent of overall diet quality, we additionally adjusted for AHEI and aMED in the primary Cox models. The core associations (F4 adverse for T2D and CKD; F7 protective for CKD; F2 protective for CAD) remained FDR-significant after adjustment for either score (Fig. 3b).

We next asked whether the factors and diet quality scores carried overlapping or distinct prognostic information using stepwise Cox models fitted in the discovery dataset and validated in the held-out replication dataset (Methods). Adding the eight NMF factors to the covariate-adjusted model improved discrimination for all three outcomes (ΔC = + 0.0018 for CAD, +0.0046 for T2D, +0.0049 for CKD; Table 1). The improvements were confirmed by bootstrapped confidence intervals and time-dependent area under the receiver operating characteristic curve (AUC) at an 8-year horizon (Table 1; Supplementary File 15). By comparison, adding AHEI or aMED individually produced smaller improvements (ΔC ≤ +0.0025). The relationship was asymmetric: adding AHEI or aMED to a model containing the NMF factors produced negligible further improvement (ΔC = ≤ +0.0005), whereas adding the factors to a model containing either score improved discrimination by +0.0017 to +0.0045 (Table 1). This indicates that the factors capture most of the diet-related prognostic signal contained in AHEI and aMED scores, while also carrying additional information that the scores do not.

**Table 1.**
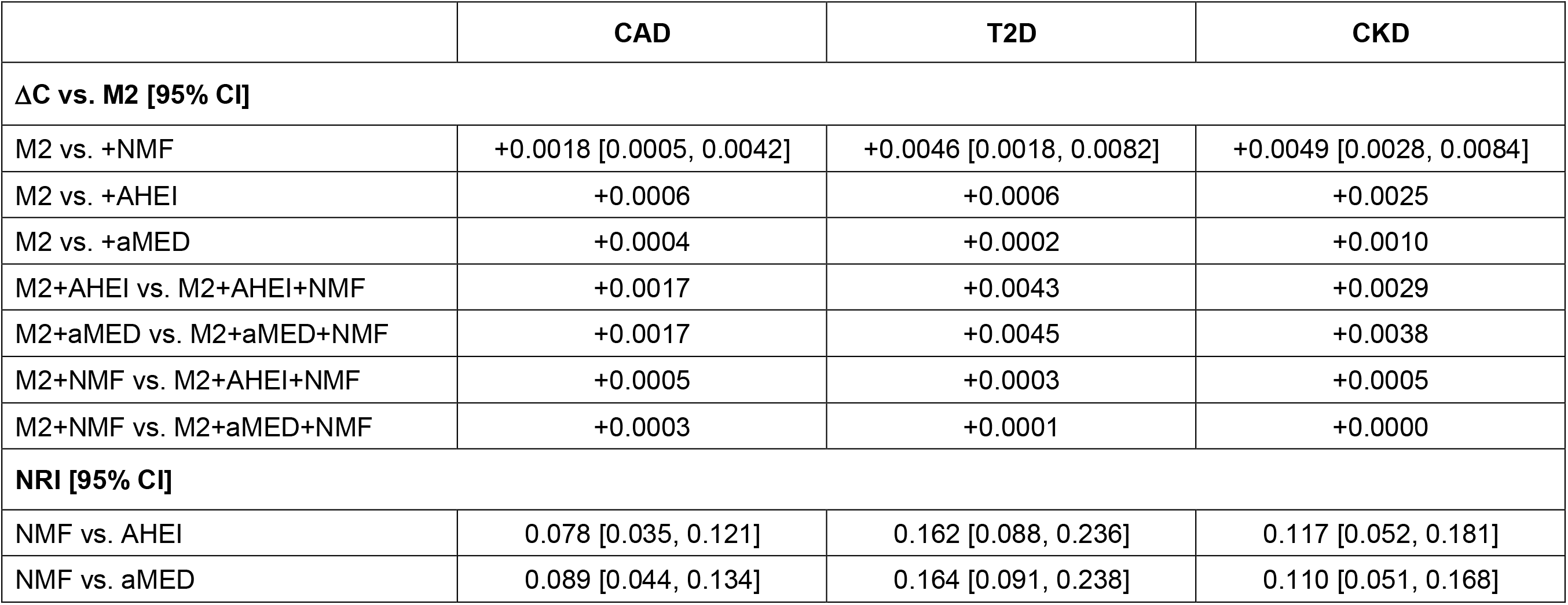
Incremental discrimination and reclassification of NMF intake factors beyond established diet quality scores in the discovery dataset. Stepwise Cox proportional hazards models were trained in the discovery dataset (*n* = 55,839) and validated in the held-out replication dataset (*n* = 23,932) using time-on-study as the timescale with M2 covariates from the primary Cox analysis. ΔC denotes the change in Harrell’s Concordance index relative to M2. Bootstrap 95% confidence intervals (CIs; 500 resamples pooled across 5 imputations via Rubin’s rules) are shown for M2 +NMF; remaining ΔC rows are point estimates computed as the difference in pooled C-indices between nested models. Category-free net reclassification improvement (NRI) was computed at an 8-year horizon; 95% CIs were obtained from 1,000 bootstrap iterations pooled across 5 imputations. AHEI, Alternative Healthy Eating Index-2010; aMED, alternate Mediterranean Diet score; NMF, values for the eight non-negative matrix factorization z-scored intake factors; CAD, coronary artery disease; T2D, type 2 diabetes; CKD, chronic kidney disease.

|  | **CAD** | **T2D** | **CKD** |
|---|---|---|---|
| **ΔC vs. M2 [95% CI]** |  |  |  |
| M2 vs. +NMF | +0.0018 [0.0005, 0.0042] | +0.0046 [0.0018, 0.0082] | +0.0049 [0.0028, 0.0084] |
| M2 vs. +AHEI | +0.0006 | +0.0006 | +0.0025 |
| M2 vs. +aMED | +0.0004 | +0.0002 | +0.0010 |
| M2+AHEI vs. M2+AHEI+NMF | +0.0017 | +0.0043 | +0.0029 |
| M2+aMED vs. M2+aMED+NMF | +0.0017 | +0.0045 | +0.0038 |
| M2+NMF vs. M2+AHEI+NMF | +0.0005 | +0.0003 | +0.0005 |
| M2+NMF vs. M2+aMED+NMF | +0.0003 | +0.0001 | +0.0000 |
| **NRI [95% CI]** |  |  |  |
| NMF vs. AHEI | 0.078 [0.035, 0.121] | 0.162 [0.088, 0.236] | 0.117 [0.052, 0.181] |
| NMF vs. aMED | 0.089 [0.044, 0.134] | 0.164 [0.091, 0.238] | 0.110 [0.051, 0.168] |

To assess whether this improved discrimination translated into better risk stratification, we computed the category-free net reclassification improvement (NRI) at an 8-year horizon. Replacing AHEI or aMED with the NMF factors improved risk classification for all three outcomes (NRI 0.08–0.16). The improvement was the largest for T2D (NRI = 0.162 [0.088, 0.236] vs. AHEI; 0.164 [0.091, 0.238] vs. aMED) and replicated in the held-out dataset (Table 1; Extended Data Table 4).

Finally, to estimate the potential population health impact, assuming causal associations, we computed population attributable fractions (PAF) for each FDR-significant factor-outcome pair (Methods). An estimated 20.9% [14.9, 27.9] of the incident CKD cases and 14.9% [7.2, 22.2] of T2D cases were attributable to the highest (vs. lowest) tertile of intake of the high sugar and processed foods factor (F4), while lowest (vs. highest) tertile of intake of the plant-based and fish (F7) and cheese and whole grains (F2) factors accounted for 17.6% [10.6, 24.9] of CKD and 8.2% [3.3, 12.7] of CAD cases, respectively (Supplementary File 17). After mutual adjustment for all eight factors, PAFs remained significant for CKD and CAD but were attenuated for T2D (Supplementary File 17).

### Intake factors have distinct metabolite profiles

To characterize their metabolic profiles, we tested the association of each factor with 249 circulating NMR metabolites (1,992 factor–metabolite pairs) in the discovery dataset and validated results in the held-out replication dataset (Methods; Fig. 1c). Across the eight factors, 1,226 pairs (61.5%) reached FDR significance (Methods), predominantly involving lipoprotein subclass remodeling and fatty acid composition. Of those, 413 (33.7%) replicated in the held-out dataset (Bonferroni-corrected *P* <0.05), consistent with prior diet–metabolite studies^20,21^ and with concordant effect sizes between datasets (Fig. 4a). Replication rates varied by factor: F4 (high sugar and processed foods) and F7 (plant-based and fish) had the highest rates (64%), whereas F3 (meat and starchy vegetables) and F5 (aromatic vegetables and rice) had few replicated associations. The metabolic profiles for F4 and F7 were consistent with their contrasting dietary compositions and cardiometabolic risk associations. F4 was associated with depletion of circulating docosahexaenoic acid (DHA; β = −0.20 per SD) and omega-3 fatty acids, whereas F7 showed strong positive associations with polyunsaturated fatty acids (PUFAs; β = +0.13) and DHA (β = +0.11). Associations with high-density lipoprotein (HDL) particle remodeling were particularly prominent for F2 (cheese and whole grains; Fig. 4b). Full MWAS results are shown in Supplementary File 19.

**Figure 4.**
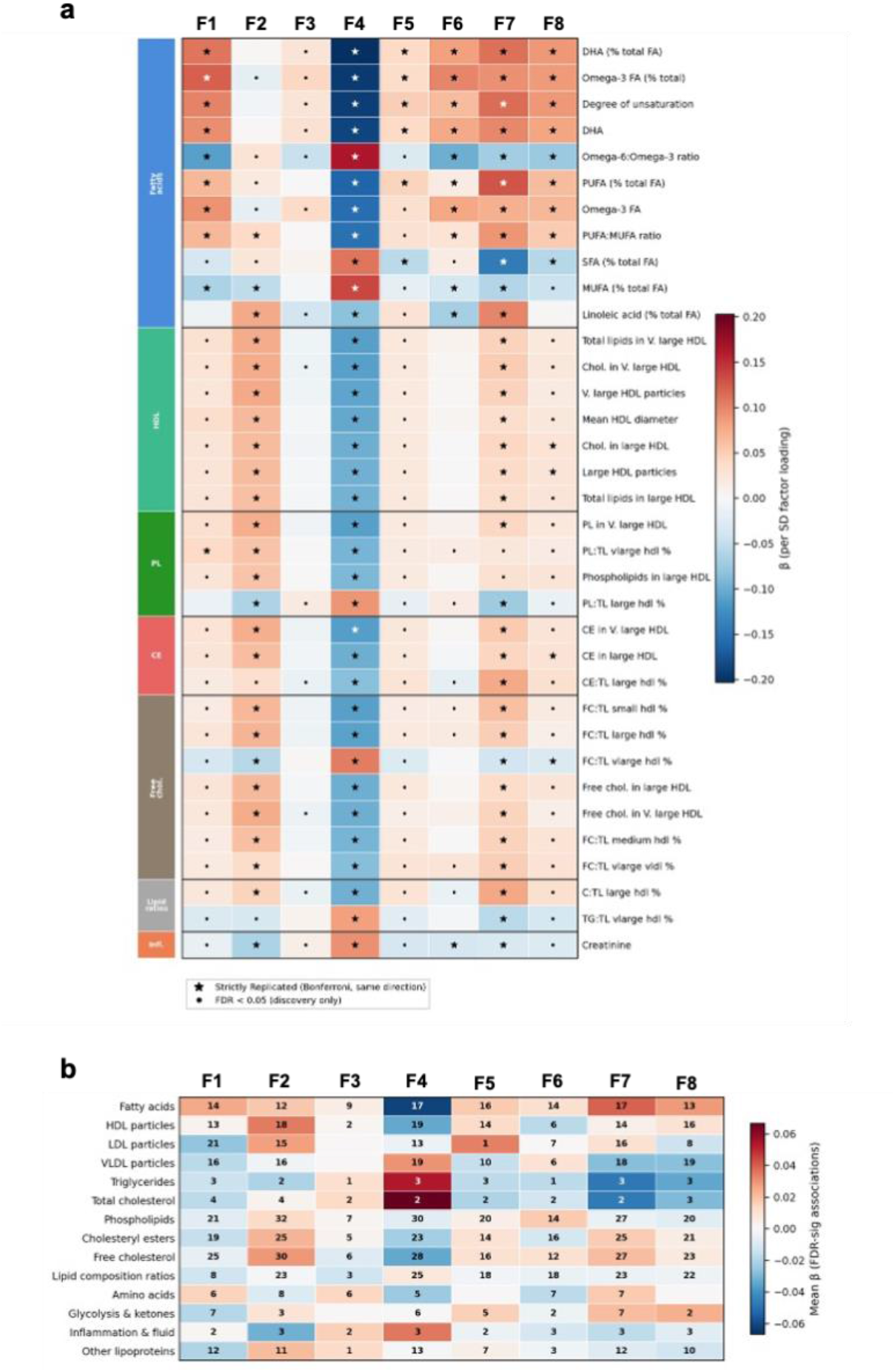
Metabolome-wide associations between NMF-derived intake factors and circulating metabolites. **a,** Heatmap of standardized regression coefficients (β per SD factor loading) from OLS regression of 249 NMR metabolites on each intake factor, adjusted for age, sex, BMI, energy intake, smoking, alcohol intake, physical activity, assessment center and fasting time (*n* = 30,063–30,508 in discovery dataset). Rows show the top-associated metabolites grouped by metabolite class. Stars indicate strict replication (Bonferroni-significant in replication dataset with concordant direction of effect); dots indicate FDR <0.05 in discovery only. **b,** Summary of FDR-significant associations per metabolite class and factor. Cell values show the number of significant associations; color intensity reflects the mean β across significant associations within each class. F1, Factor 1 (prudent base); F2, Factor 2 (cheese and whole grains); F3, Factor 3 (meat and starchy vegetables); F4, Factor 4 (high sugar and processed); F5, Factor 5 (aromatic vegetables and rice); F6, Factor 6 (fruit and low-fat dairy); F7, Factor 7 (plant-based and fish); F8, Factor 8 (raw salad); HDL, high-density lipoprotein; PL, phospholipids; CE, cholesterol esters; FC, free cholesterol; Infl., inflammatory; DHA, docosahexaenoic acid; FA, fatty acid; PUFA, polyunsaturated fatty acid; MUFA, monounsaturated fatty acid; TG, triglycerides; TL, total lipid; LDL, low-density lipoprotein; VLDL, very low-density lipoprotein.

### Participants with the same diet quality scores but different intake profiles differ in cardiometabolic risk markers

To test whether the additional information captured by NMF manifests in clinical biomarker profiles, we matched participants in the replication dataset (*n* = 23,932) on their diet quality score (energy-adjusted AHEI or aMED) and total energy intake, using a caliper of 0.25 SD on each variable (Methods; Fig. 1c). Matching on energy intake ensured that differences between paired participants reflected dietary composition rather than how much they ate overall, yielding 11,958 AHEI-matched and 11,959 aMED-matched pairs. We then computed an NMF-derived composite cardiometabolic risk score for each participant (Methods) and designated the higher-scoring member of each pair as the “adverse” member and the other as the “favorable” member. Among the top 5% of AHEI-matched pairs with the most divergent NMF profiles, termed discordant pairs (*n* = 598), adverse members had significantly higher BMI (Cohen’s *d* = 0.22, FDR *q* = 0.006), body fat percentage (*d* = 0.19, *q* = 0.006), white blood cell count (*d* = 0.26, *q* = 0.001) and lower vitamin D (*d* = −0.20, *q* = 0.008; Fig. 5a, Supplementary Table 13). Among aMED-discordant pairs, differences extended to lipids (HDL-C: *d* = −0.25; triglycerides: *d* = 0.14; ApoA: *d* = −0.19; all *q* < 0.05) and inflammatory markers (CRP: *d* = 0.19, *q* = 0.012; Fig. 5b, Supplementary Table 14). Thus, people whom diet quality scores rate as equally healthy can differ meaningfully in what they eat and in clinical risk markers.

**Figure 5.**
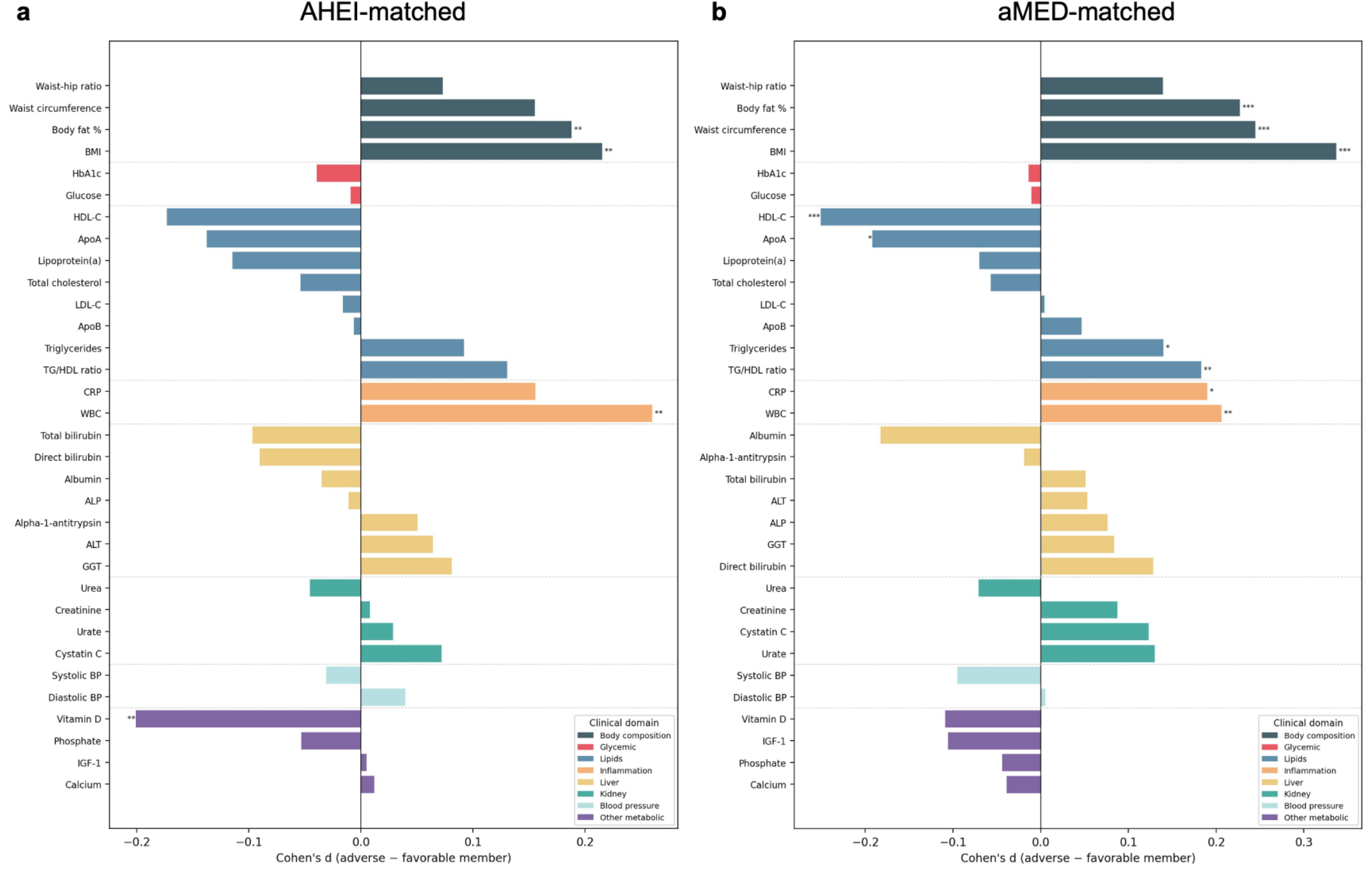
Biomarker differences between diet score-matched participants with discordant NMF profiles. **(a)** Horizontal bar plot showing differences in 33 clinical biomarkers between adverse and favorable members among the top 5% most NMF-discordant AHEI-matched pairs (*n* = 598) in the replication dataset. Discordance was defined by cosine distance of W-matrix vectors. Participants were matched 1:1 on energy-adjusted AHEI score and total energy intake (caliper 0.25 SD). Within each extreme-discordant pair, members were classified as adverse or favorable according to an NMF-derived composite cardiometabolic risk score. Bars represent Cohen’s d effect sizes (adverse minus favorable member) categorized across clinical domains. **(b)** Same analysis for aMED-matched pairs (*n* = 598). In both panels, significance from Wilcoxon signed-rank tests with Benjamini-Hochberg FDR correction is indicated by asterisks (* *q*<0.05, ** *q*<0.01, *** *q*<0.001). BMI, body mass index; HbA1c, hemoglobin A1c; HDL-C, high density lipoprotein cholesterol; ApoA, apolipoprotein A; LDL-C, low-density lipoprotein cholesterol; ApoB, apolipoprotein B; TG/HDL, triglyceride to HDL ratio; CRP, C-reactive protein; WBC, white blood cell count; ALT, alanine transaminase; ALP, alkaline phosphatase; GGT, gamma-glutamyl transferase; BP, blood pressure; IGF-1, insulin-like growth factor 1.

To illustrate risk attribution at the individual level, we examined one of the most discordant pairs, matched not only on AHEI score (∼41) but also on sex, age, BMI, smoking status and alcohol intake (Methods; Extended Data Fig. 2a). Individual A’s diet was dominated by F1 (prudent base; 64%), with smaller contributions from F4 (high sugar and processed foods; 17%) and F2 (cheese and whole grains; 14%). Individual B’s diet was more evenly spread, led by F5 (aromatic vegetables and rice; 41%), followed by F8 (raw salad; 15%), F2 (14%) and F7 (plant-based and fish; 13%; Extended Data Fig 2b). Despite similar AHEI scores, Individual A’s profile predicted higher CKD risk (relative HR = 1.21, computed from that individual’s factor scores relative to the cohort mean) and Individual B’s higher T2D risk (relative HR = 1.24), each driven by different foods (Extended Data Fig. 2c). For T2D, Individual A’s risk was driven mainly by crisps, white bread, added sugar, sweets and fried food, whereas Individual B’s came from refined grains and red meat. Dietary advice for Individual A might therefore focus on reducing sugary and processed snacks, and for Individual B on replacing refined grains and red meat with whole grains and poultry, seafood or plant-based protein.

## Discussion

Applying NMF to food-level intake data from 87,162 UKB participants, we identified eight reproducible intake factors with four significant factor-disease associations: the high sugar and processed foods factor (F4) with higher risk of T2D and CKD, and the cheese and whole grains (F2) and plant-based and fish (F7) factors with lower risk of CAD and CKD, respectively. The factors had distinct metabolic profiles, with F4 showing a pro-atherogenic profile and F7 a strong shift toward PUFAs. The factors added prognostic value beyond both the AHEI and aMED, whereas neither score added meaningfully beyond the factors, and among with matched scores, those with the most divergent factor profiles differed in adiposity, inflammatory and lipid biomarkers. Food-level intake factors therefore capture clinically relevant dietary variation that diet quality scores mask.

The factor-disease associations identified here refine findings from prior dietary pattern studies. The adverse F4 factor recapitulates the “Western” pattern linked to T2D and cardiovascular risk in PCA-based studies^22–25^. The association with CKD may partly reflect the high sodium content of several top-weighted F4 foods and ultra-processed foods in general, consistent with evidence linking sodium intake to reduced kidney function^26,27^. The protective F7 factor aligns with foods emphasized by AHEI and aMED^28–31^ and the “prudent” pattern that has been linked to healthier outcomes^23,32,33^. A notable finding was that the coffee, cheese and whole grains factor (F2) was the only factor protective for CAD. This is consistent with evidence supporting coffee^34–36^ and full-fat and fermented dairy^37–39^ as protective for cardiometabolic disease. The separation of cheese into a distinct factor from other dairy products would likely be diluted if dairy intake were aggregated into a single group, illustrating the value of food-level resolution. Although not significant after multiple testing correction, the nominally adverse association between the aromatic vegetables and rice factor (F5) and T2D also highlights this advantage. While vegetable intake is generally considered protective, the vegetables in this factor are co-consumed with high-glycemic white rice and white pasta, which may offset their benefits for blood sugar control. Our approach differs from prior NMF applications^17,18,40^ in three key aspects: we retained 148 individual foods rather than aggregating to food groups, we preserved continuous mixture representations rather than clustering participants into discrete groups and we tested whether NMF-derived factors carried prognostic information beyond established diet quality scores.

A central finding of this work is that NMF intake factors add prognostic value beyond established diet quality scores, and that the reverse is not true. When both were tested in stepwise Cox regression models, the factors subsumed nearly all diet-related prognostic signal captured by AHEI and aMED, while retaining significant discrimination and reclassification gains. This asymmetry likely reflects a fundamental difference in how dietary intake is represented. Whereas AHEI and aMED compress food intake into a single score^11,12^, NMF preserves the multidimensional structure of dietary intake behavior. The discrimination advantage replicated for T2D and CKD in the held-out dataset; the attenuation for CAD likely reflects limited statistical power (∼970 events), though the direction of improvement was consistent. The PAF estimates further suggest that the identified factors could represent meaningful targets for cardiometabolic disease prevention, with the largest attributable burdens observed for CKD. After mutual adjustment for all eight factors, the independent PAFs remained significant for CKD and CAD, whereas the T2D PAF for F4 was attenuated, suggesting that part of this factor’s population-level T2D burden is shared with correlated intake factors.

These findings have practical implications at three levels. For research, food-level factors complement rather than replace diet quality scores. The scores remain simple, guideline-based summaries that are easy to communicate, whereas the factors capture variation the scores miss, including two factors (F3 and F8) that were largely uncorrelated with either score, and they identify which combinations of foods carry risk. For clinical care, an individual’s factor profile could direct counseling to the specific foods driving their predicted risk, as the matched-pair example illustrates, rather than to an overall score that two people can reach through very different diets. For population health, if the associations are causal, the PAF estimates suggest that shifting intake away from the high sugar and processed foods factor could prevent a meaningful share of incident T2D and CKD. Because the gains in discrimination were modest in absolute terms, the factors are better viewed as a way to target dietary advice than as a stand alone risk prediction tool.

The metabolomics results provide insights into the biological characteristics associated with each factor and their potential relevance to cardiometabolic disease. The depletion of circulating DHA, omega-3 fatty acids and elevated very-low-density lipoprotein triglycerides characterizing the high sugar and processed foods factor closely mirrors lipidomic biomarkers of ultra-processed diets and low seafood intake^41^. This likely reflects displacement of nutrient-rich foods by energy-dense foods^27^. Conversely, the enrichment in PUFAs for the plant-based and fish factor aligns with controlled feeding studies that identify circulating PUFAs as biomarkers of nut and plant-based oil consumption^42^. The shift toward large, cholesterol-rich HDL particles associated with the cheese and whole grains factor is consistent with metabolomic studies linking dairy and whole grain intake to favorable lipoprotein profiles^43^. These divergent profiles suggest that distinct eating behaviors influence cardiometabolic disease through heterogeneous biological mechanisms that a single summary score cannot resolve.

The study should be interpreted in the light of some limitations. The factor structure replicated with high fidelity within individuals recruited in England but showed attenuation in those recruited in Scotland and Wales, suggesting that dietary factors are shaped by population-level food cultures and would need to be re-derived in cohorts with substantially different dietary norms. Additionally, although NMF is data-driven, analytical decisions including food preprocessing and the choice of eight factors involve subjective judgement. Finally, NMF introduces eight continuous exposures compared with a single diet quality score, raising the question of overfitting. However, likelihood ratio tests penalize for additional degrees of freedom, the adjusted R^2^ metric accounts for model complexity and the factors added discrimination beyond models already containing AHEI or aMED while the reverse did not hold.

Despite these limitations, the UKB’s scale, linked EHR records and the discovery-replication-validation design^53^ allowed us to derive and replicate intake factors, test their associations with cardiometabolic disease, characterize their metabolic signatures and identify individuals whose cardiometabolic risk is masked by diet quality scores. The food-level resolution of individual risk profiles could enable more targeted dietary counseling by identifying which foods drive a given patient’s cardiometabolic risk. Integrating NMF-based profiling into clinical workflows would require validation in diverse populations and development of interpretable reporting tools.

These results suggest several directions for future work, including validation in diverse cohorts with different dietary cultures and assessment instruments, randomized trials testing whether targeting an individual’s adverse factors improves cardiometabolic risk beyond standard guidance, and extensions that capture meal timing and interactions between foods, which additive factors cannot represent^57,59^. Ultimately, our findings demonstrate that food-level dietary intake profiling captures clinically meaningful variation that established diet quality scores mask, offering a path toward more personalized dietary assessment to support cardiometabolic health.

## Methods

### Ethical Approval

UK Biobank (UKB) data access was approved by the UKB through project application number 1251. The UKB study received ethical approval from the NHS National Research Ethics Service (Ref 11/NW/0382), and all participants provided informed consent. Researchers do not need separate approval.

### Study population

The UKB is a multi-center prospective cohort study of more than 500,000 participants aged 40– 70 years at enrollment^44^. Participants were recruited from 2006 to 2010 at 22 assessment centers across England, Scotland and Wales, with baseline information, health and lifestyle questionnaires, physical measures and biological samples collected according to standardized procedures^45–47^. Between 2009 and 2012 a subset of participants completed up to five online 24-hour dietary recall assessments using the Oxford WebQ^48^. We restricted the analysis to participants who completed at least two valid dietary recalls to better reflect usual dietary intake ^28^, resulting in an initial sample of 102,316 participants. The Oxford WebQ has demonstrated acceptable validity and reproducibility when using ≥2 dietary recalls^49^. A dietary recall was considered invalid if it reported atypical intake, duration of less than 5 minutes, had no completion time or reported implausible daily energy intake, defined as <800 or >4,200 kcal/d for male and <500 or >3,500 kcal/d for female^28,50^ participants. We excluded participants who were pregnant, or had prevalent CAD, T2D, CKD, other diabetes mellitus or cancer. Cancer was defined using the union of 11 cancer sites with non-melanoma skin cancer (ICD-10 C44) excluded^51^. Participants with glycated hemoglobin (HbA1c) ≥6.0% (42 mmol/mol) were excluded to remove individuals with undiagnosed or borderline diabetes^52^. We defined baseline as the date of each participant’s last valid dietary recall. The final analytic cohort included 87,162 participants.

For analyses requiring independent cohorts, we used a three-way geographic split^51^. The discovery (*n*=55,839) and replication (*n*=23,932) datasets included participants recruited from assessment centers in England using a 70/30 random split stratified by age, sex and number of valid dietary recalls. The validation dataset (*n*=7,391) included all participants recruited from Scotland and Wales, providing an independent test of generalizability across regional dietary cultures. The discovery and replication datasets were recombined (*n*=79,771) for all primary Cox association analyses after structure replication of dietary factors was confirmed. The discovery dataset was retained separately for metabolomics association analysis, added prognostic value model training and discordance analysis; the replication dataset served as the held-out validation cohort for each of these analyses. The Scotland and Wales validation dataset was held out exclusively for NMF factor generalization assessment^51^.

### Dietary assessment

Dietary intake was assessed using the validated Oxford WebQ, a web-based 24-hour recall dietary assessment tool^48^. Out of the 102,316 participants with at least two valid dietary recalls, 42,898 completed two, 28,884 completed three, 13,511 completed four and 1,869 completed five. Participants reported their consumption of up to 206 food and beverage items during the previous day on each occasion. For this study, we derived intake estimates in servings per day, averaging intake across all valid recalls for each participant to approximate usual dietary exposure. 196 individual foods and beverages remained after recoding and quality control (Supplementary Information).

#### Food matrix construction

We removed alcoholic beverages and plain drinking water from the food matrix prior to factor derivation. Alcohol was excluded from the NMF input matrix because alcohol intake (continuous grams (g) per day) was included separately as a covariate in the outcome models, avoiding overlap between the derived dietary factors and the adjustment variables^51^. Plain drinking water was excluded because it contributed no nutrients and loaded heavily across all NMF-derived factors in preliminary analyses, diluting factor-specific signal. Rarely consumed foods and beverages (<5% of population) were removed. The remaining 148 food intake values were preprocessed in three sequential steps: winsorization at the 99 ^th^ percentile, log transformation (log(1 + x)) and division by the standard deviation (SD) of each food. Preprocessing parameters including the rarely consumed foods that were dropped, 99 ^th^ percentile thresholds and standard deviations, were computed on the discovery dataset and applied to the replication and validation datasets.

#### Diet quality scores

Estimated nutrients and energy intake from the Oxford WebQ were used to compute two *a priori* diet quality scores, the Alternative Healthy Eating Index-2010 (AHEI) and the alternate Mediterranean Diet Score (aMED). For each participant, the mean intakes across all valid 24-hour dietary recalls were analyzed. The AHEI score was developed based on the consumption of 11 foods and nutrients that have been shown to predict chronic disease risk, including fruits, vegetables, nuts and legumes, alcohol, red and processed meat, whole grains, sodium, trans fats and long-chain omega(ω)-3 fatty acids^53^. For this study, we used the AHEI-2010 version, which has been adapted to the UKB^11^. We could not include the energy from trans-fat due to a lack of information in the UKB, and we used seafood intake for the ω-3 component because the UKB only provides information about total ω-3 intake^31^. Each component was scored continuously from 0 (least healthy) to 10 (most healthy), producing a total score ranging from 0 to 100 (Supplementary Table 1). The aMED score calculates adherence to 9 eating behaviors that characterize the Mediterranean diet^12^. Intakes of vegetables, legumes, fruits, nuts, whole grains, fish, and monounsaturated (MUFA) to saturated (SFA) fatty acid ratio above the median received 1 point, while intakes below the median received 0 points. Intakes of red and processed meats above the median received 0 points, and intakes below the median received 1 point (Supplementary Table 2). Alcohol intake between 5–25g received 1 point, with all other intake levels receiving 0 points. The final score ranged from 0 to 9. For both AHEI and aMED, a higher score reflects overall better diet quality^11,12^. Both scores were adjusted for total energy intake using the Willett residual method, in which the raw score is regressed on total energy intake (kcal/day) and the residual is added to the score predicted at the sample mean energy intake ^54^. Total energy intake included energy from alcohol and energy-adjusted scores were used in all downstream analyses.

### Outcome ascertainment

Incident events for CAD, T2D and CKD were identified using the UKB first occurrence of health outcomes data (Category 1712), which maps Read codes from primary care, ICD-9 and ICD-10 codes from hospital inpatient data, and ICD-10 codes from death registrations to three-character ICD-10 codes with the first occurrence date as the date the code was first recorded across any source^55^. Incident events were defined using registry-grade evidence only (hospital inpatient records, death registrations or primary care) with participants flagged as incident by self-report alone treated as censored^51^. First occurrences data were accessed on 22 September 2022, with censoring dates of 30 September 2021, 31 July 2021, and 31 October 2021 for participants recruited in England, Scotland and Wales, respectively.

Baseline prevalence for all diseases and clinical risk factors was assessed using a combination of first occurrence data, cancer registry data, self-reported illness, medication use, surgical history and/or biochemical markers. Baseline prevalent CAD, T2D, CKD, other diabetes mellitus and cancer cases were excluded from the cohort. Incident cases of other or unspecified diabetes mellitus (n=473) were additionally excluded to control for reverse causation^56^. Participants were followed from the date of their last valid dietary recall until the first occurrence of CAD, T2D or CKD, death, loss to follow-up or the end of outcome ascertainment based on the nation-specific censor dates, whichever came first. Data sources, ICD codes and evidence rules for all prevalent and incident disease definitions are provided in Supplementary Table 5.

### Assessment of covariates

Multivariable models were adjusted for a range of demographic, socioeconomic, lifestyle and clinical covariates that were selected at baseline based on the existing literature. Covariates were organized into three sequential tiers of adjustment. Model 1 (M1, minimally-adjusted) included total energy intake (quintiles). Model 2 (M2, primary) additionally included ethnicity (White or non-White), highest educational qualification (7, 10, 13, 15, 19 or 20 years), annual household income (<18k, 18-31k, 31-52k, 52-100k or >100k), smoking status (never, former or current), alcohol intake (g/day), leisure-time physical activity (low, moderate or high), body mass index (BMI, kg/m^2^), dietary supplement use (yes/no), family history of diabetes (yes/no) and family history of heart disease (yes/no). Nation was not included as a covariate because datasets were split by nation. The Cox analyses were sex-stratified, rather than including sex as a covariate, and age was the timescale (see Cox regression). Additional details on derived variable calculation and quality control can be found in Supplementary Information.

### Missing data imputation

In contrast to data with genuine missingness, a “Prefer not to answer” (PNA) response can be considered an informative non-response^57^. PNA responses were therefore retained as an explicit factor level for annual household income, smoking status, ethnicity, highest educational qualification and supplement use, rather than recoding as missing. The average percentage of missing data across all final variables was ∼1.0% (range: 0–4%; Supplementary Table 6). We performed missing data imputation separately for each dataset (discovery, replication and validation) using multiple imputation by chained equations (MICE) with the R package mice^58^. For each, five multiply imputed datasets were generated with a maximum of ten iterations for each imputation. The variables used as predictors in the imputation model included all baseline covariates, NMF intake factor shares, energy-adjusted diet quality scores (AHEI, aMED), and the Nelson-Aalen estimate of cumulative hazard and event indicator for each primary outcome. All statistical analyses were run independently in each of the five imputed datasets, with results pooled using Rubin’s rule^59,60^.

### Statistical analysis

All statistical analyses were performed in R v4.6.1 and Python v3.11.

#### Non-negative matrix factorization

We applied NMF to the preprocessed food intake matrix V (participants X food) composed of the intake of 148 foods and beverages (servings/d) across 55,839 individuals in the discovery dataset. NMF decomposes the input matrix into two lower-rank non-negative matrices: a participant-by-factor loading matrix W and a factor-by-food weight matrix H, such that V ≍ W x H^19^. NMF summarizes the information available in food intake data into an optimal number of *k* latent intake factors and for each factor, each food has a weight that represents the contribution of the food to the construction of that factor^17^. The non-negativity constraint of NMF ensures that every factor is interpretable as an additive combination of foods consumed^19^. NMF was implemented using scikit-learn v1.6.1 NMF class with coordinate descent solver, Frobenius loss and α_H = 0.005 and l1_ratio = 0.80 as regularization parameters.

##### Rank selection

The number of latent factors (rank *k*) was determined through a multi-phase selection process involving stability analyses on the discovery dataset and evaluation of factor reproducibility in the replication dataset. In the first phase, explained variance, residual sum of squares and sparsity indices were computed using deterministic NNDSVD initialization^61^ across ranks *k* = 2–40 without regularization to identify a candidate range. In the second phase, the sensitivity of factor structure to L1 regularization was evaluated across a grid of α_H (0.0–0.5) and l1_ratio (0.8, 1.0) values. Factor structure was largely insensitive to penalty strength, reflecting the intrinsic sparsity of the food intake matrix (79.7% zeros). In the third phase, factor stability was assessed at candidate ranks *k* = 8, 10, 12, 14 using 30 random initializations per rank, with stability evaluated by Hungarian-matched H matrix cosine similarity across all pairwise run comparisons and 10 split-half resamples. In the fourth phase, the representative factorization was selected from 30 random initializations as the run with the lowest reconstruction error, following a multi-start approach analogous to that used in latent factor decomposition of genetic data^62^. We selected *k* = 8 as the final rank based on de novo replication performance (see below); all eight factors replicated with cosine similarity >0.98 between independently fitted discovery and replication models, whereas = 10 had two factors with cosine similarity <0.65 that failed to replicate. At *k* = 8, the factorization explained 29.2% of the squared Frobenius norm in the food intake matrix. Full rank selection diagnostics are provided in Supplementary Fig. 1–4.

##### Pattern interpretation

Factors were interpreted based on H-matrix food loadings. For each factor, the relative contribution (%) of each food was calculated as the ratio of that food’s weight to the sum of all food weights within the factor^18^. Foods with a relative contribution ≥ 2.5% were considered active in each factor^18^. An exclusivity index was calculated as the ratio of each food’s loading on a given factor to its total loading across all factors, distinguishing foods unique to a given factor from widely consumed staples. Labels were assigned descriptively from the active foods in each factor.

##### Archetype assignment

To descriptively characterize individual-level factor dominance, each participant’s factor loadings were normalized to proportional contributions by dividing by their row sum. The archetype threshold was determined by starting at 50% and lowering in 10% increments until each archetype contained at least 200 participants, yielding a final threshold of 40%. Participants whose highest proportional loading exceeded this threshold were assigned to that factor’s archetype; all others were classified as Mixed. The raw salad (F8) archetype contained too few participants (n=18) for meaningful characterization and was merged into the Mixed group.

##### De novo replication

To confirm that the factors reflected stable dietary structure rather than sample-specific noise, we fitted NMF independently in the discovery, replication and validation datasets using identical hyperparameters (*k* = 8, α_H = 0.005, l1_ratio = 0.80, 50 random initializations, 500 maximum iterations). The representative factorization was selected for each dataset as the run with the lowest reconstruction error. Factors were aligned across dataset pairs using the Hungarian algorithm applied to the pairwise cosine similarity matric of H-matrix rows. Per-factor cosine similarity, Spearman ρ on food loadings and Jaccard overlap of the top 10 foods were computed for each matched factor pair.

##### Generalization

To assess the generalizability of the factors, the H-matrix was frozen from the discovery dataset and held-out participants were projected onto it using non-negative least squares (NNLS) via scikit-learn’s NMF.transform(). Given held-out data *V^test^* and fixed H, *W^test^* = min‖*V^test^* − *W^test^H*‖^2^, subject to *W^test^* ≥ 0. Generalization was evaluated in the replication (within-population) and validation (geographically independent) datasets by comparing reconstruction performance with that of the discovery dataset and examining differences in per-factor loading distributions using Kolmogorov-Smirnov tests.

#### Cox proportional hazards regression

Cox proportional hazards models were used to estimate hazard ratios (HR) and 95% confidence intervals (CIs) for the association between each NMF factor and incident cardiometabolic disease. Analyses were conducted in the combined English sample (discovery + replication)^51^. Given the strong confounding effect of age on both dietary intake and disease risk, we used age as the timescale. Participants entered the risk set at their age at baseline (date of last valid dietary recall) and exited at their age at event or censoring^63^. Events occurring within the first year of follow-were excluded to reduce the potential influence of reverse causation. The baseline hazard was stratified via sex with ties handled using the Efron approximation.

Factor scores (W-matrix loadings) were z-score standardized using the combined-sample mean and SD for each factor and all eight factors were entered into each model simultaneously, such that the HR for a given factor represents the change in hazard per 1-SD increase, holding all other factors constant. Three tiers of covariate adjustment (M1, M2, M3) were applied as described above, with M2 as the primary model. Results were pooled across *m* = 5 imputed datasets using Rubin’s rule for coefficient estimates and the D_2_ statistic for likelihood ratio tests^59,60^. Dose-response gradients were examined by categorizing participants into those with zero loading on a given factor and tertiles of positive factor loading (T1 – T3) with the zero-loading group as the reference. E values were computed for significant HRs to assess the potential impact of unmeasured confounding^64,65^. Pre-specified subgroup analyses tested for effect modification by sex, age (above/below median), BMI (<25 vs. ≥25 kg/m^2^), physical activity, smoking status and ethnicity (White vs. non-White). Multiple comparisons were corrected using Benjamini-Hochberg false discovery rate (FDR) within each outcome (eight tests per outcome); FDR-corrected *P* < 0.05 was considered statistically significant. The proportional hazards assumption was assessed using Schoenfeld residuals and visual inspection of scaled residual plots, which revealed no clinically meaningful time-varying trends (Supplementary File 8).

##### Sensitivity analyses

Three pre-specified sensitivity analyses assessed the robustness of the primary Cox associations. First, the washout period was extended to 2 years to further address reverse causation. Second, analyses were restricted to participants with three or more valid dietary recalls to assess robustness when dietary assessment was more complete (n = 44,264). Third, BMI was removed from the M2 covariate set to quantify the proportion of the association potentially mediated through adiposity. As a sensitivity analysis for the factorization itself, we repeated NMF using Sparse NMF (SNMF/R^66^) with the Python package nimfa^67^ with an explicit sparsity penalty on the H-matrix. Full sensitivity analysis results are provided in Supplementary Table 8).

##### Compositional data analysis

Dietary intake is inherently compositional; increasing consumption of one food necessarily displaces another when total energy intake is held constant^68^. To address this constraint, we applied log-ratio transformations that enable valid inference about the substitution structure of dietary intake^69^. The primary compositional analysis used the additive log-ratio (ALR) transformation^70^. For each participant, *k* – 1 = 7 log-ratios 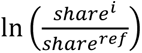 were computed, with the factor that was the most prevalent dominant loading in the population (F1) as the reference^70^. The HR for a given ALR term represents the risk change associated with substituting dietary share from the reference factor toward the alternative factor. Factors with loading weights of zero have undefined log-ratios; zeros were handled using multiplicative replacement.

##### Added prognostic value

To quantify the incremental prognostic value of NMF factors beyond established diet quality scores, we compared six stepwise Cox proportional hazards models, following the approach of Argentieri *et al*^51^: M2 (primary covariates only), M2 + NMF (all eight z-scored NMF factors), M2 + AHEI (energy-adjusted AHEI), M2 + aMED (energy-adjusted aMED), M2 + NMF + AHEI and M2 + NMF + aMED. AHEI and aMED were always entered in separate models. Models were trained on the discovery dataset (*n* = 55,839) and validated on the held-out replication dataset (*n* = 23,932). For this analysis we used time-on-study as the timescale with age at baseline as covariate. Explained variation was quantified using *R*^2^ from the CoxR2 package^71^. Discrimination was quantified using Harrell’s C-index with bootstrapped 95% confidence intervals on ΔC versus M2 (500 resamples). Both *R*^2^ and C-index were pooled across the five MICE imputations via Fisher’s z transformation: *R*^2^ values were transformed by taking the square root, converted to z-scale, averaged, inverse-transformed, and then squared^51,72^. C-index values were mapped to Somers’ D (D = 2C – 1) before Fisher’s z transformation and mapped back after averaging. Likelihood ratio tests comparing each expanded model to M2 were pooled via the *D*^2^ statistic.

Time-dependent area under the receiver operating characteristic curve (tdAUC) was computed at 8-year horizons using the timeROC package^73^ and pooled across imputations via Fisher’s z transformation and Somers’ D, as above. Category-free net reclassification improvement (NRI) was computed at an 8-year time horizon using the nricens package^74,75^. Linear predictors were converted to absolute predicted risks at the NRI horizon via the stratum-specific baseline cumulative hazard before computing reclassification. NRI was computed for each of the five MICE-imputed datasets and pooled via Rubin’s rules. Confidence intervals were obtained from 1,000 bootstrap iterations within nricens.

##### Population attributable fraction

For each FDR-significant factor–outcome association from the primary Cox regression, we estimated the population attributable fraction (PAF) using tertile contrasts. For adverse associations, the PAF was defined as the fraction of cases among individuals in the highest exposure tertile (T3) that would be prevented if their risk were reduced to that of the lowest tertile (T1); for protective associations, the direction was reversed. The unadjusted PAF used Levin’s formula with the crude population prevalence of exposure^76^. The mutually adjusted PAF, which held the other seven NMF factor loadings constant, used Miettinen’s formula with exposure prevalence among cases^77^, as applying multivariable-adjusted HR in Levin’s formula with the crude population prevalence produces biased estimates^76,78^. Bootstrap 95% confidence intervals were obtained from 500 resamples.

##### Random survival forest

To rank the relative importance of NMF factors for each outcome and to explore potential non-linear associations, we fitted random survival forests (RSF) separately for each primary outcome using the R package randomForestSRC^79^ on the discovery dataset. As randomForestSRC implements missingness handling at each node split, RSF models used the pre-imputation dataset rather than multiply imputed datasets. Missing data rates were low (maximum 4.0% for leisure-time physical activity with all other covariates <1.6%). The exposure variables were the eight NMF-derived factor loadings entered as continuous, unstandardized W-matrix scores. Covariates matched the M2 Cox model specification except for total energy intake, which was entered as a continuous variable rather than quintile dummies^80^. Default hyperparameter values were used (ntree = 1,000, nodesize = 15, importance = “permute”). Model performance was assessed using out-of-bag (OOB) prediction error, computed as 1 minus the OOB concordance index and convergence was confirmed by visual inspection of OOB error versus number of trees. Two complementary variable ranking methods were computed for each outcome: permutation variable importance (VIMP)^81^ and minimal depth^82^.

#### Diet quality score discordance analysis

To investigate whether NMF factors capture dietary variation not fully represented by conventional diet quality scores, we identified individuals with closely matched AHEI or aMED scores but maximally divergent NMF factor loadings. All discordance analyses were conducted in the replication dataset (n = 23,932) using Cox regression coefficients estimated on the discovery dataset only.

For each diet quality score (AHEI and aMED, analyzed separately), 1:1 greedy nearest-neighbor matching without replacement was performed on energy-adjusted diet score and mean total energy intake using a caliper of 0.25 SD on each dimension^83^. Discordance between score-matched pairs was quantified using cosine distance on L1-normalized W vectors. Extreme-discordant pairs were defined as those in the top 5% of cosine distance. Within each extreme-discordant pair, members were classified as “adverse” or “favorable” using a composite risk score:

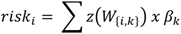

where *z*(*W*_{_*_i_*_,*k*}_) is the z-scored factor loading and *β_k_* is the mean log(HR) for factor *k* across CAD, T2D and CKD from the discovery-only M2 Cox model. The pair member with the higher composite risk score was labeled “adverse.” Pair members were compared across 33 cardiometabolic risk factors using Cohen’s d and Wilcoxon signed-rank tests. Benjamini-Hochberg FDR correction was applied across all 33 biomarker tests. To quantify the percentage of individuals classified as having high diet quality have elevated cardiometabolic risk when assessed by NMF factors, we computed composite risk scores among individuals in the top decile of AHEI and aMED and reported the fraction with above-median risk relative to the full dataset.

##### Individual risk heterogeneity

To illustrate the clinical relevance of NMF factors at the individual level, we selected a single pair from the discordance analysis for detailed illustration. Within the top 10% most NMF-discordant pairs, candidates were filtered to same-sex pairs and scored by *selection score* = *cosine distance* − (*λ x covariate distance*), where covariate distance is the mean standardized absolute difference across energy intake, alcohol intake, smoking status, age and BMI. This soft-matching approach balanced maximal NMF discordance against covariate matching. To identify which individual foods contribute most to each participant’s NMF-derived risk, we computed outcome-specific food harm scores:

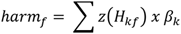

where *H_kf_* is the food’s z-scored weight on factor *k* from the discovery H-matrix and *β_k_* is the outcome-specific coefficient from the discovery-only Cox analysis. Each participant’s food-level risk was then computed as *intake_f_x* ℎ*arm_f_* where *intake_f_* is their mean reported intake (servings/day). Clinical interpretation of food-level differences between case study participants was provided by a registered dietitian on the study team.

#### NMF metabolomics association analysis

To characterize the metabolic profiles associated with the NMF-derived intake factors, we conducted a metabolome-wide association study (MWAS) using nuclear magnetic resonance (NMR) spectroscopy metabolomics from the UKB Nightingale Health platform^84^. A total of 249 metabolites were measured in plasma samples collected at baseline (instance 0), including 168 directly measured and 81 derived ratios. Metabolites with more than 50% missing values were excluded. Not all participants in the analytic cohort had NMR metabolomics data available. The sample sizes were *n* = 30,063 – 30,508 in the discovery dataset and *n* = 23,932 in the replication dataset, depending on metabolite-level missingness. Identical analytical procedures were applied independently within each cohort.

Metabolite concentrations were inverse-normal transformed and NMF factor loadings were z-score standardized, applied within each dataset separately. For each NMF factor independently, we fitted ordinary least squares (OLS) regression models for each metabolite: metabolite ∼ factor + covariates. Covariates included the Cox regression M2 covariate set (age at baseline, sex, BMI, total energy intake, alcohol intake, ethnicity, smoking status, educational attainment, household income, leisure-time physical activity and dietary supplement use) plus UKB assessment center (22 collection sites as a categorical variable) and fasting time (hours since last meal). Multiple testing was corrected using Benjamini-Hochberg FDR applied per factor (249 tests per factor). FDR-corrected *P* <0.05 was considered statistically significant. Significant associations in the discovery dataset were tested in the replication dataset. An association was considered replicated if *P* <0.05 with the same direction of effect. Beta concordance between discovery and replication datasets was assessed using Pearson correlation of effect estimates.

## Reporting summary

Further information on research design is available in the Nature Portfolio Reporting Summary linked to this article.

## Data Availability

UK Biobank data are available to approved researchers through the UK Biobank Access Management System (https://www.ukbiobank.ac.uk). This study was conducted under application number 1251. Source data for the main figures are provided with this paper.

## Code Availability

Code is available at https://github.com/hdkittrell/nmf.

## Extended Data

**Extended Data Table 1.** Baseline descriptive statistics according to NMF-derived intake factor archetype.

| **NMF Archetype** | **Prudent base (F1)** | **Cheese & whole grains (F2)** | **Meat & starchy vegetables (F3)** | **High sugar & processed (F4)** | **Aromatic vegetables & rice (F5)** |
|---|---|---|---|---|---|
| Participants, n (%) | 25,193 (28.9%) | 9,833 (11.3%) | 1,854 (2.1%) | 3,114 (3.6%) | 420 (0.5%) |
| Age at baseline | 59.3 (7.7) | 60.1 (7.7) | 59.6 (7.9) | 55.0 (7.7) | 55.0 (7.5) |
| Female | 52.9% | 46.2% | 56.2% | 29.2% | 66.2% |
| **Ethnicity** |  |  |  |  |  |
| White | 96.6% | 98.5% | 99.4% | 95.6% | 91.0% |
| Non-White | 3.1% | 1.1% | 0.6% | 4.3% | 8.6% |
| Income >100k | 5.0% | 10.6% | 4.0% | 4.2% | 8.6% |
| Years of education | 16.3 (4.5) | 17.8 (3.7) | 14.6 (4.9) | 15.1 (4.7) | 16.9 (4.2) |
| Family history of diabetes, yes | 17.1% | 14.0% | 17.0% | 19.6% | 23.3% |
| Family history of CVD, yes | 44.1% | 42.2% | 43.5% | 38.6% | 41.9% |
| Prevalent HTN | 48.2% | 45.9% | 55.9% | 50.5% | 47.6% |
| Prevalent Dyslipidemia | 47.2% | 49.8% | 52.6% | 52.0% | 45.0% |
| Supplement use, yes | 33.2% | 27.4% | 26.4% | 24.2% | 36.0% |
| BMI, kg/m<sup>2</sup> | 26.2 (4.2) | 25.9 (4.1) | 27.3 (4.4) | 28.1 (4.8) | 26.8 (4.8) |
| Never smoker | 61.9% | 59.9% | 50.2% | 53.3% | 53.3% |
| Physical activity, high | 34.0% | 32.3% | 32.7% | 31.7% | 36.4% |
| Alcohol intake, g/day | 11.7 (15.9) | 19.8 (20.0) | 19.6 (23.1) | 17.7 (24.0) | 24.2 (26.4) |
| Energy intake, kcal/day | 1,949 (454) | 2,125 (462) | 1,986 (491) | 2,245 (547) | 1,915 (476) |
| Protein, g/day | 78.8 (19.1) | 78.7 (18.4) | 82.3 (19.8) | 80.0 (22.3) | 75.4 (22.2) |
| Fat, g/day | 64.7 (21.4) | 80.1 (23.8) | 70.5 (23.3) | 84.0 (25.9) | 65.1 (23.0) |
| Carbohydrates, g/day | 255.4 (64.8) | 251.0 (63.0) | 233.2 (67.1) | 275.7 (74.3) | 226.1 (63.7) |
| Fiber, g/day | 18.7 (5.6) | 17.1 (5.0) | 17.8 (5.5) | 12.6 (4.2) | 18.3 (6.6) |
| Saturated Fat, g/day | 23.5 (9.1) | 32.1 (10.7) | 26.2 (10.0) | 31.8 (11.4) | 22.5 (9.5) |
| AHEI score (energy-adjusted) | 49.8 (11.9) | 44.0 (11.2) | 44.0 (10.9) | 32.5 (10.1) | 50.2 (11.9) |
| aMED score (energy-adjusted) | 4.3 (1.7) | 3.3 (1.6) | 3.3 (1.5) | 2.2 (1.3) | 4.2 (1.5) |
| **NMF Archetype** | **Fruit & LF dairy (F6)** | **Plant-based & fish (F7)** | **Mixed** | **Full Cohort** |  |
| Participants, n | 700 (0.8%) | 902 (1.0%) | 45,146 (51.8%) | 87,162 |  |
| Age at baseline | 58.6 (7.5) | 57.6 (8.2) | 59.3 (7.8) | 59.2 (7.8) |  |
| Female | 81.3% | 78.4% | 62.8% | 57.0% |  |
| **Ethnicity** |  |  |  |  |  |
| White | 94.9% | 92.7% | 96.9% | 96.9% |  |
| Non-White | 4.8% | 6.9% | 2.8% | 2.8% |  |
| Income >100k | 6.4% | 10.0% | 7.8% | 7.1% |  |
| Years of education | 15.7 (4.6) | 17.6 (3.7) | 16.7 (4.3) | 16.6 (4.4) |  |
| Family history of diabetes, yes | 20.1% | 18.0% | 17.2% | 16.9% |  |
| Family history of CVD, yes | 46.9% | 44.3% | 44.0% | 43.6% |  |
| Prevalent HTN | 54.1% | 33.4% | 47.6% | 47.8% |  |
| Prevalent Dyslipidemia | 50.3% | 35.1% | 49.0% | 48.6% |  |
| Supplement use, yes | 39.6% | 55.8% | 34.1% | 32.8% |  |
| BMI, kg/m<sup>2</sup> | 28.2 (5.4) | 24.4 (4.0) | 26.3 (4.4) | 26.3 (4.3) |  |
| Never smoker | 51.9% | 60.0% | 58.0% | 58.9% |  |
| Physical activity, high | 36.2% | 34.5% | 34.0% | 33.7% |  |
| Alcohol intake, g/day | 10.8 (16.3) | 10.9 (15.7) | 15.7 (18.4) | 15.1 (18.5) |  |
| Energy intake, kcal | 1,704 (439) | 1,984 (478) | 2,043 (465) | 2,027 (471) |  |
| Protein, g/day | 73.8 (22.8) | 75.0 (22.4) | 80.9 (19.4) | 79.9 (19.4) |  |
| Fat, g/day | 53.2 (19.4) | 83.3 (26.0) | 73.2 (23.2) | 71.7 (23.6) |  |
| Carbohydrates, g/day | 224.7 (69.4) | 226.9 (70.0) | 250.6 (65.5) | 252.0 (65.8) |  |
| Fiber, g/day | 16.6 (6.4) | 21.6 (7.1) | 18.7 (5.9) | 18.3 (5.8) |  |
| Saturated Fat, g/day | 19.2 (7.9) | 24.3 (9.4) | 26.8 (9.8) | 26.5 (10.2) |  |
| AHEI score (energy-adjusted) | 49.6 (11.6) | 62.4 (11.4) | 49.9 (12.5) | 48.6 (12.7) |  |
| aMED score (energy-adjusted) | 4.2 (1.5) | 5.9 (1.5) | 4.2 (1.8) | 4.0 (1.8) |  |

Baseline demographic, lifestyle and clinical characteristics of 87,162 UK Biobank participants, stratified by NMF-derived intake factor archetype. Participants whose dominant factor accounted for more than 40% of their total factor loadings were assigned to that factor’s archetype; all others were classified as Mixed. Values are mean (SD) for continuous variables and % for categorical variables. CVD, cardiovascular disease; HTN, hypertension; g/day, grams per day; kcal, kilocalories; AHEI, Alternative Healthy Eating Index; aMED, Alternate Mediterranean Diet score; CAD, coronary artery disease; T2D, type 2 diabetes; CKD, chronic kidney disease.

**Extended Data Table 2.** Explained variation and C-index across multivariable models in the discovery dataset.

| **Disease** | **Model 1** |  | **Model 2** |  | **Model 3** |  | **Model 4** |  | **Model 5** |  | **Model 6** |  | **Cases** | **Sample** |
|---|---|---|---|---|---|---|---|---|---|---|---|---|---|---|
|  | *C-index* | *R<sup>2</sup>* | *C-index* | *R<sup>2</sup>* | *C-index* | *R<sup>2</sup>* | *C-index* | *R<sup>2</sup>* | *C-index* | *R<sup>2</sup>* | *C-index* | *R<sup>2</sup>* |  |  |
| CAD | 0.687 | 0.354 | 0.689 | 0.358 | 0.688 | 0.355 | 0.688 | 0.355 | 0.689 | 0.360 | 0.689 | 0.359 | 2,270 | 55,477 |
| T2D | 0.793 | 0.677 | 0.797 | 0.693 | 0.793 | 0.682 | 0.793 | 0.681 | 0.797 | 0.694 | 0.797 | 0.693 | 991 | 55,607 |
| CKD | 0.744 | 0.545 | 0.748 | 0.567 | 0.746 | 0.556 | 0.745 | 0.551 | 0.749 | 0.569 | 0.748 | 0.567 | 1,143 | 55,560 |

Model 1: M1, primary Cox model 2 (M2) covariates (age, sex, energy intake, ethnicity, education, income level, smoking status, alcohol intake, physical activity, BMI, family history of heart disease, family history of diabetes, supplement use). Model 2: M1 + eight NMF factor scores. Model 3: M1+ AHEI. Model 4: M1 + aMED. Model 5: M1 + NMF + AHEI. Model 6: M1 + NMF + aMED. Models were fit in the discovery dataset (England, *n* = 55,839) using Cox proportional hazards regression with time-on-study as the timescale. Cases and sample sizes shown are from the M2 model after exclusion of participants with missing data. CAD, coronary artery disease; T2D, type 2 diabetes; CKD, chronic kidney disease; C-index, Harrell's concordance statistic; R<sup>2</sup>, Cox-Snell R<sup>2</sup> from CoxR2 package.

**Extended Data Table 3.** Explained variation and C-index across multivariable models in the replication dataset.

| **Disease** | **Model 1** |  | **Model 2** |  | **Model 3** |  | **Model 4** |  | **Model 5** |  | **Model 6** |  | **Cases** | **Sample** |
|---|---|---|---|---|---|---|---|---|---|---|---|---|---|---|
|  | *C-index* | *R<sup>2</sup>* | *C-index* | *R<sup>2</sup>* | *C-index* | *R<sup>2</sup>* | *C-index* | *R<sup>2</sup>* | *C-index* | *R<sup>2</sup>* | *C-index* | *R<sup>2</sup>* |  |  |
| CAD | 0.689 | 0.355 | 0.689 | 0.357 | 0.689 | 0.355 | 0.689 | 0.354 | 0.689 | 0.355 | 0.689 | 0.355 | 947 | 23,771 |
| T2D | 0.775 | 0.608 | 0.779 | 0.620 | 0.779 | 0.618 | 0.776 | 0.609 | 0.780 | 0.622 | 0.778 | 0.618 | 356 | 23,839 |
| CKD | 0.749 | 0.565 | 0.752 | 0.575 | 0.751 | 0.572 | 0.749 | 0.569 | 0.751 | 0.572 | 0.752 | 0.574 | 508 | 23,825 |

Models were trained in the discovery dataset and validated by applying the fitted linear predictor to the held-out replication dataset (*n* = 23,932). Model definitions as in Extended Data Table 2. Cases and sample sizes shown are from the M2 model after exclusion of participants with missing data. CAD, coronary artery disease; T2D, type 2 diabetes; CKD, chronic kidney disease; C-index, Harrell’s concordance statistic; R<sup>2</sup>, Cox-Snell R<sup>2</sup> from CoxR2 package.

**Extended Data Table 4.**
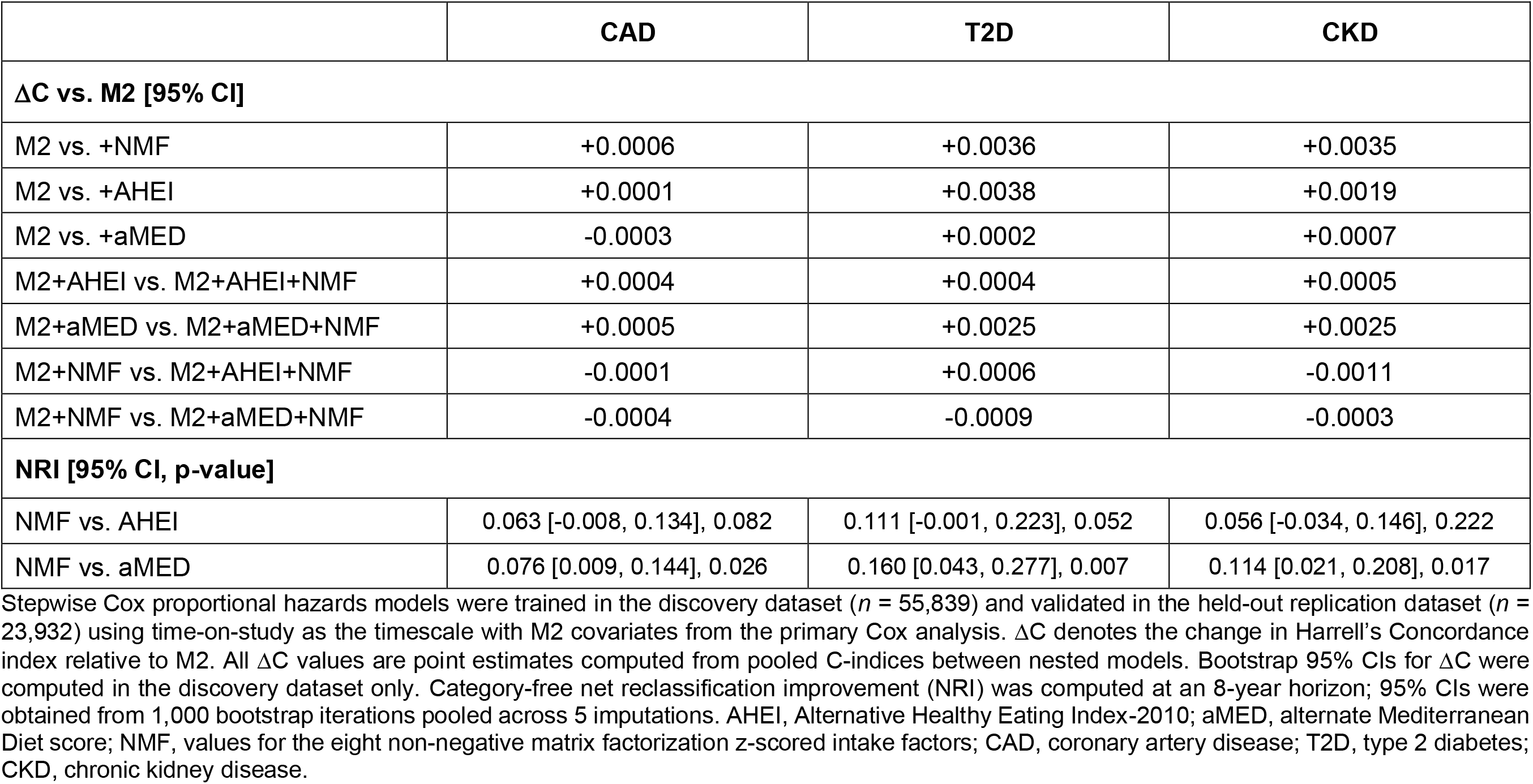
Incremental discrimination and reclassification of NMF intake factors beyond established diet quality scores in the replication dataset.

**Extended Data Figure 1.**
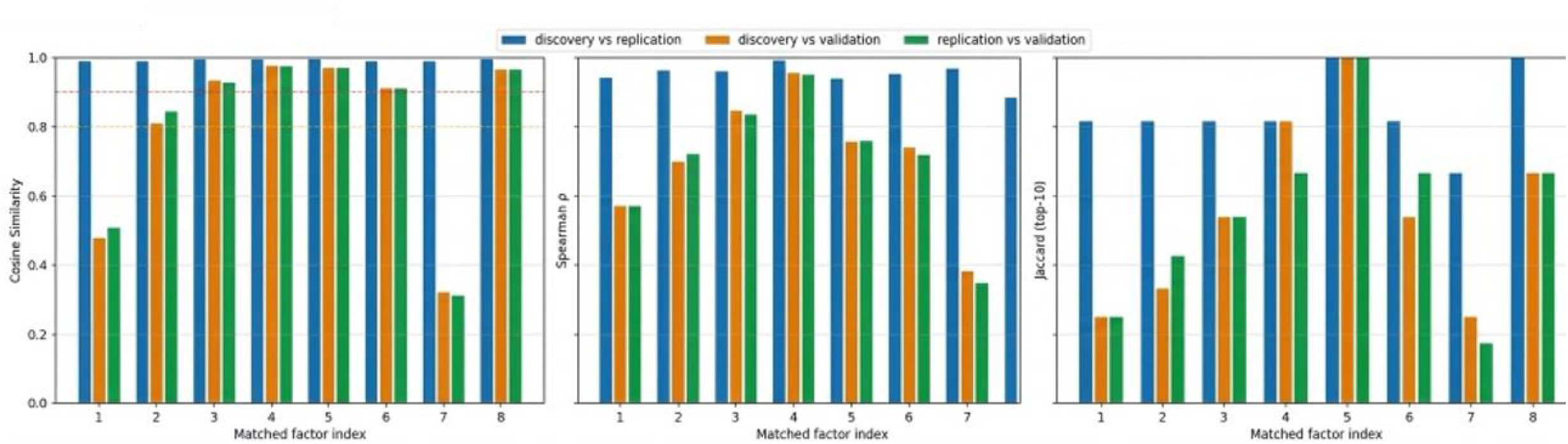
De novo replication of NMF intake factors across datasets. Independent NMF decompositions (*k* = 8) were performed separately in the discovery (*n* = 55,839), replication (*n* = 23,932) and validation (*n* = 7,931) datasets using the same preprocessed food intake matrix and hyperparameters. Factors were matched across datasets by maximum cosine similarity of H-matrix loading vectors. Three complementary metrics are shown for each matched factor pair: cosine similarity of full loading vectors (**left**), Spearman rank correlation of food loadings (**middle**) and Jaccard index of the top-10 highest loading foods (**right**).

**Extended Data Figure 2.**
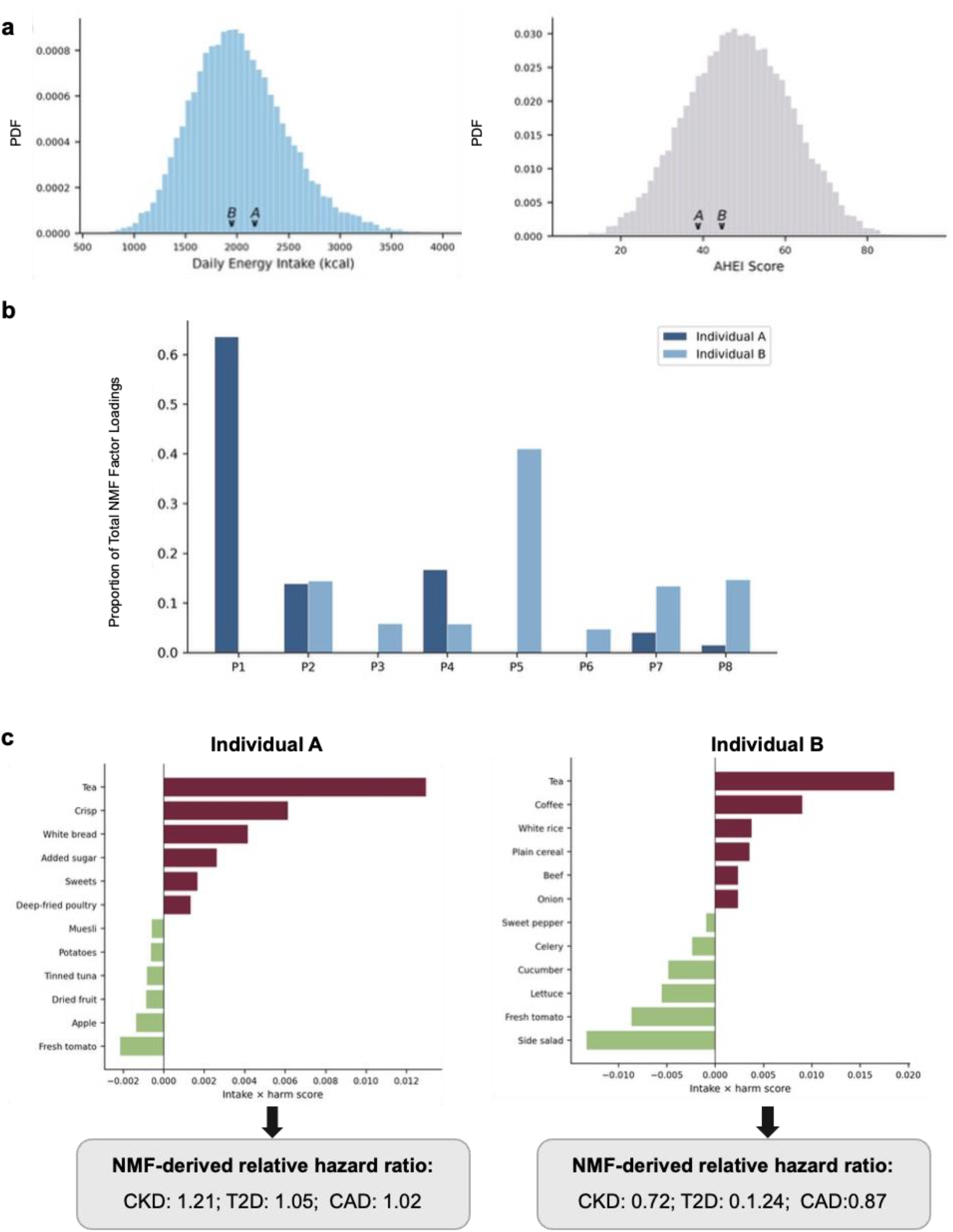
Food-level risk attribution in two participants with similar AHEI scores. **(a)** Probability density function histograms of daily caloric intake (kcal) and Alternative Healthy Eating Index-2010 (AHEI) scores showing that the two selected individuals A and B have similar caloric intake (∼2,000 kcal) and AHEI scores (∼41.0). The individuals are both same-sex participants of similar age, BMI, smoking status, and alcohol intake. Arrows denote the positions of Individual A and Individual B within the cohort distributions. **(b)** Histograms showing proportional distribution of NMF factor loadings (F1-F8) for Individual A (dark blue) and Individual B (light blue). **(c)** Bar plots showing food contributions to overall cardiometabolic risk (calculated as intake X harm score) for Individual A (left) and Individual B (right). Risk-contributing foods are shown in red, and protective foods in green. The NMF-derived relative hazard ratios per outcome are shown below each bar plot. The relative hazards were derived by applying Cox regression coefficients derived in the discovery dataset to individual factor loadings of pair matched in replication dataset. PDF, probability density function; NMF, non-negative matrix factorization; CAD, coronary artery disease; CKD, chronic kidney disease; T2D, type 2 diabetes.

## References

1. Naghavi, M. et al. Global burden of 292 causes of death in 204 countries and territories and 660 subnational locations, 1990–2023: a systematic analysis for the Global Burden of Disease Study 2023. The Lancet 406, 1811–1872 (2025).

2. Brauer, M. et al. Global burden and strength of evidence for 88 risk factors in 204 countries and 811 subnational locations, 1990–2021: a systematic analysis for the Global Burden of Disease Study 2021. The Lancet 403, 2162–2203 (2024).

3. Willett, W. et al. Food in the Anthropocene: the EAT–Lancet Commission on healthy diets from sustainable food systems. The Lancet 393, 447–492 (2019).

4. Afshin, A. et al. Health effects of dietary risks in 195 countries, 1990–2017: a systematic analysis for the Global Burden of Disease Study 2017. The Lancet 393, 1958–1972 (2019).

5. Leme, A. C. B., Hou, S., Fisberg, R. M., Fisberg, M. & Haines, J. Adherence to Food-Based Dietary Guidelines: A Systemic Review of High-Income and Low- and Middle-Income Countries. Nutrients 13, 1038 (2021).

6. Krebs-Smith, S. M., Guenther, P. M., Subar, A. F., Kirkpatrick, S. I. & Dodd, K. W. Americans Do Not Meet Federal Dietary Recommendations. The Journal of Nutrition 140, 1832–1838 (2010).

7. Klapp, A.-L., Simorangkir, J. P., Gallani, V., Braune, D. A. B. & Risius, A. From Policy to Practice: A Global Analysis to Identify Factors That Influence the Implementation of Food-Based Dietary Guidelines by Health and Nutrition Professionals. Current Developments in Nutrition 10, 109509 (2026).

8. Lee, B. Y. et al. Research gaps and opportunities in precision nutrition: an NIH workshop report. The American Journal of Clinical Nutrition 116, 1877–1900 (2022).

9. Guasch-Ferré, M. et al. Precision nutrition for cardiometabolic diseases. Nat Med 31, 1444–1453 (2025).

10. Lee, B. Y. et al. Research gaps and opportunities in precision nutrition: an NIH workshop report. The American Journal of Clinical Nutrition 116, 1877–1900 (2022).

11. Chiuve, S. E. et al. Alternative dietary indices both strongly predict risk of chronic disease. J Nutr 142, 1009–1018 (2012).

12. Trichopoulou, A., Costacou, T., Bamia, C. & Trichopoulos, D. Adherence to a Mediterranean diet and survival in a Greek population. N Engl J Med 348, 2599–2608 (2003).

13. Merino, J. et al. Polygenic scores, diet quality, and type 2 diabetes risk: An observational study among 35,759 adults from 3 US cohorts. PLoS Med 19, e1003972 (2022).

14. Zhao, J. et al. A review of statistical methods for dietary pattern analysis. Nutrition Journal 20, 37 (2021).

15. Wingrove, K., Lawrence, M. A. & McNaughton, S. A. A Systematic Review of the Methods Used to Assess and Report Dietary Patterns. Front. Nutr. 9, 892351 (2022).

16. Schulz, C.-A., Oluwagbemigun, K. & Nöthlings, U. Advances in dietary pattern analysis in nutritional epidemiology. Eur J Nutr 60, 4115–4130 (2021).

17. Zetlaoui, M., Feinberg, M., Verger, P. & Clémençon, S. Extraction of Food Consumption Systems by Nonnegative Matrix Factorization (NMF) for the Assessment of Food Choices. Biometrics 67, 1647–1658 (2011).

18. Gazan, R. et al. Dietary patterns in the French adult population: a study from the second French national cross-sectional dietary survey (INCA2) (2006–2007). Br J Nutr 116, 300–315 (2016).

19. Lee, D. D. & Seung, H. S. Learning the parts of objects by non-negative matrix factorization. Nature 401, 788–791 (1999).

20. Long, M. et al. Plasma metabolomic signature of a proinflammatory diet in relation to breast cancer risk: a prospective cohort study. The American Journal of Clinical Nutrition 121, 548–557 (2025).

21. Li, J. et al. The Mediterranean diet, plasma metabolome, and cardiovascular disease risk. European Heart Journal 41, 2645–2656 (2020).

22. Joshi, S., Kalantar-Zadeh, K., Chauveau, P. & Carrero, J. J. Risks and Benefits of Different Dietary Patterns in CKD. American Journal of Kidney Diseases 81, 352–360 (2023).

23. Van Dam, R. M., Rimm, E. B., Willett, W. C., Stampfer, M. J. & Hu, F. B. Dietary Patterns and Risk for Type 2 Diabetes Mellitus in U.S. Men. Ann Intern Med 136, 201–209 (2002).

24. Nettleton, J. A., Steffen, L. M., Ni, H., Liu, K. & Jacobs, D. R. Dietary Patterns and Risk of Incident Type 2 Diabetes in the Multi-Ethnic Study of Atherosclerosis (MESA). Diabetes Care 31, 1777–1782 (2008).

25. Guallar-Castillón, P. et al. Major dietary patterns and risk of coronary heart disease in middle-aged persons from a Mediterranean country: The EPIC-Spain cohort study. *Nutrition*, Metabolism and Cardiovascular Diseases 22, 192–199 (2012).

26. McMahon, E. J. et al. A randomized trial of dietary sodium restriction in CKD. J Am Soc Nephrol 24, 2096–2103 (2013).

27. Menichetti, G., Ravandi, B., Mozaffarian, D. & Barabási, A.-L. Machine learning prediction of the degree of food processing. Nat Commun 14, 2312 (2023).

28. Chen, S.-H., Liu, B., Cheng, H.-M., Liang, L. & Hu, F. B. Dietary Patterns and Risk of Cardiovascular Disease and All-Cause Mortality in Adults with Untreated or Uncontrolled Hypertension. The Journal of Nutrition 156, 101659 (2026).

29. Wang, P. et al. Optimal dietary patterns for prevention of chronic disease. Nat Med 29, 719–728 (2023).

30. Tessier, A.-J. et al. Optimal dietary patterns for healthy aging. Nat Med 31, 1644–1652 (2025).

31. Maroto-Rodriguez, J. et al. Diet quality patterns and chronic kidney disease incidence: a UK Biobank cohort study. The American Journal of Clinical Nutrition 121, 445–453 (2025).

32. Hu, F. B. et al. Prospective study of major dietary patterns and risk of coronary heart disease in men. The American Journal of Clinical Nutrition 72, 912–921 (2000).

33. Heidemann, C. et al. Dietary Patterns and Risk of Mortality From Cardiovascular Disease, Cancer, and All Causes in a Prospective Cohort of Women. Circulation 118, 230–237 (2008).

34. Lu, X. et al. Habitual Coffee, Tea, and Caffeine Consumption, Circulating Metabolites, and the Risk of Cardiometabolic Multimorbidity. The Journal of Clinical Endocrinology & Metabolism 110, e1845–e1855 (2025).

35. O’Keefe, J. H. et al. Effects of Habitual Coffee Consumption on Cardiometabolic Disease, Cardiovascular Health, and All-Cause Mortality. Journal of the American College of Cardiology 62, 1043–1051 (2013).

36. Marcus, G. M. et al. Caffeine and Cardiovascular Disease: A Scientific Statement From the American Heart Association. Circulation 154, (2026).

37. Hirahatake, K. M. et al. Potential Cardiometabolic Health Benefits of Full-Fat Dairy: The Evidence Base. Advances in Nutrition 11, 533–547 (2020).

38. Astrup, A., Geiker, N. R. W. & Magkos, F. Effects of Full-Fat and Fermented Dairy Products on Cardiometabolic Disease: Food Is More Than the Sum of Its Parts. Advances in Nutrition 10, 924S–930S (2019).

39. Aune, D., Norat, T., Romundstad, P. & Vatten, L. J. Dairy products and the risk of type 2 diabetes: a systematic review and dose-response meta-analysis of cohort studies. The American Journal of Clinical Nutrition 98, 1066–1083 (2013).

40. De Gavelle, E., Huneau, J.-F. & Mariotti, F. Patterns of Protein Food Intake Are Associated with Nutrient Adequacy in the General French Adult Population. Nutrients 10, 226 (2018).

41. Zhao, L. et al. Metabolomic and Proteomic Signatures of Ultra-processed Foods Are Positively Associated with Adverse Liver Outcomes. The Journal of Nutrition 155, 1851–1858 (2025).

42. Guertin, K. A. et al. Metabolomics in nutritional epidemiology: identifying metabolites associated with diet and quantifying their potential to uncover diet-disease relations in populations. The American Journal of Clinical Nutrition 100, 208–217 (2014).

43. Feeney, E. L. et al. Dairy matrix effects: response to consumption of dairy fat differs when eaten within the cheese matrix—a randomized controlled trial. The American Journal of Clinical Nutrition 108, 667–674 (2018).

44. UK Biobank. UK Biobank: a protocol for a large-scale prospective epidemiological resource [Internet]. (2007).

45. Bycroft, C. et al. The UK Biobank resource with deep phenotyping and genomic data. Nature 562, 203–209 (2018).

46. Elliott, P., Peakman, T. C., & on behalf of UK Biobank. The UK Biobank sample handling and storage protocol for the collection, processing and archiving of human blood and urine. International Journal of Epidemiology 37, 234–244 (2008).

47. Sudlow, C. et al. UK Biobank: An Open Access Resource for Identifying the Causes of a Wide Range of Complex Diseases of Middle and Old Age. PLoS Med 12, e1001779 (2015).

48. Liu, B. et al. Development and evaluation of the Oxford WebQ, a low-cost, web-based method for assessment of previous 24 h dietary intakes in large-scale prospective studies. Public Health Nutr. 14, 1998–2005 (2011).

49. Carter, J. L. et al. Reproducibility of dietary intakes of macronutrients, specific food groups, and dietary patterns in 211 050 adults in the UK Biobank study. J Nutr Sci 8, e34 (2019).

50. Cole, J. B., Westerman, K. E., Manning, A. K., Florez, J. C. & Hirschhorn, J. N. Genetic heritability as a tool to evaluate the precision of 24-hour recall dietary questionnaire variables in UK Biobank. Front. Genet. 13, 1070511 (2023).

51. Argentieri, M. A. et al. Integrating the environmental and genetic architectures of aging and mortality. Nat Med 31, 1016–1025 (2025).

52. Dorans, K. S. et al. Effects of a Low-Carbohydrate Dietary Intervention on Hemoglobin A_1c_: A Randomized Clinical Trial. JAMA Netw Open 5, e2238645 (2022).

53. McCullough, M. L. et al. Diet quality and major chronic disease risk in men and women: moving toward improved dietary guidance. Am J Clin Nutr 76, 1261–1271 (2002).

54. Willett, W. & Stampfer, M. J. TOTAL ENERGY INTAKE: IMPLICATIONS FOR EPIDEMIOLOGIC ANALYSES. American Journal of Epidemiology 124, 17–27 (1986).

55. UK Biobank. UK Biobank: First occurrence of health outcomes defined by 3-character ICD10 code [internet]. (2019).

56. Zhao, C. et al. Associations of Combined Genetic and Lifestyle Risks With Incident Type 2 Diabetes in the UK Biobank. Diabetes 75, 860–866 (2026).

57. Thomas, D. M. et al. Machine learning modeling practices to support the principles of AI and ethics in nutrition research. Nutr. Diabetes 12, 48 (2022).

58. Van Buuren, S. & Groothuis-Oudshoorn, K. mice: Multivariate Imputation by Chained Equations. 3.19.0 10.32614/CRAN.package.mice (2006).

59. Marshall, A., Altman, D. G., Holder, R. L. & Royston, P. Combining estimates of interest in prognostic modelling studies after multiple imputation: current practice and guidelines. BMC Med Res Methodol 9, 57 (2009).

60. Li, K. H., Raghunathan, T. E. & Rubin, D. B. Large-Sample Significance Levels from Multiply Imputed Data Using Moment-Based Statistics and an *F* Reference Distribution. Journal of the American Statistical Association 86, 1065–1073 (1991).

61. Boutsidis, C. & Gallopoulos, E. SVD based initialization: A head start for nonnegative matrix factorization. Pattern Recognition 41, 1350–1362 (2008).

62. Morze, J. et al. Decomposing polygenic risk of coronary artery disease into distinct biological processes. Preprint at 10.64898/2026.10.08.26365088 (2026).

63. Thiébaut, A. C. M. & Bénichou, J. Choice of time-scale in Cox’s model analysis of epidemiologic cohort data: a simulation study. Statistics in Medicine 23, 3803–3820 (2004).

64. VanderWeele, T. J. & Ding, P. Sensitivity Analysis in Observational Research: Introducing the E-Value. Ann Intern Med 167, 268–274 (2017).

65. Tessier, A.-J. et al. Optimal dietary patterns for healthy aging. Nat Med 31, 1644–1652 (2025).

66. Kim, H. & Park, H. Sparse non-negative matrix factorizations via alternating non-negativity-constrained least squares for microarray data analysis. Bioinformatics 23, 1495–1502 (2007).

67. Zitnik, M. & Zupan, B. Nimfa: A Python Library for Nonnegative Matrix Factorization. Journal of Machine Learning Research 13, 849–853 (2012).

68. Ibsen, D. B. et al. Food substitution models for nutritional epidemiology. The American Journal of Clinical Nutrition 113, 294–303 (2021).

69. Aitchison, J. The Statistical Analysis of Compositional Data. Journal of the Royal Statistical Society: Series B (Methodological) 44, 139–177 (1982).

70. Greenacre, M., Martínez-Álvaro, M. & Blasco, A. Compositional Data Analysis of Microbiome and Any-Omics Datasets: A Validation of the Additive Logratio Transformation. Front. Microbiol. 12, 727398 (2021).

71. You, H. & Xu, R. CoxR2: R-squared measure based on partial LR statistic, for the Cox PH regression mode. (2022).

72. Harel, O. The estimation of *R*^2^ and adjusted *R*^2^ in incomplete data sets using multiple imputation. Journal of Applied Statistics 36, 1109–1118 (2009).

73. Blanche, P., Dartigues, J. & Jacqmin-Gadda, H. Estimating and comparing time-dependent areas under receiver operating characteristic curves for censored event times with competing risks. Statistics in Medicine 32, 5381–5397 (2013).

74. Inoue, E. nricens: NRI for Risk Prediction Models with Time to Event and Binary Response Data. (2018).

75. Pencina, M. J., D’Agostino, R. B. & Steyerberg, E. W. Extensions of net reclassification improvement calculations to measure usefulness of new biomarkers. Statistics in Medicine 30, 11–21 (2011).

76. Ferguson, J., Alvarez, A., Mulligan, M., Judge, C. & O’Donnell, M. Bias assessment and correction for Levin’s population attributable fraction in the presence of confounding. Eur J Epidemiol 39, 111–119 (2024).

77. Miettinen, O. S. PROPORTION OF DISEASE CAUSED OR PREVENTED BY A GIVEN EXPOSURE, TRAIT OR INTERVENTION1. American Journal of Epidemiology 99, 325–332 (1974).

78. Rockhill, B., Newman, B. & Weinberg, C. Use and misuse of population attributable fractions. Am J Public Health 88, 15–19 (1998).

79. Ishwaran, H. & Kogalur, U. B. Fast Unified Random Forests for Survival, Regression, and Classification (RF-SRC). (2026).

80. Baralou, V., Kalpourtzi, N. & Touloumi, G. Individual risk prediction: Comparing random forests with Cox proportional-hazards model by a simulation study. Biometrical J 65, 2100380 (2023).

81. Ishwaran, H. & Lu, M. Standard errors and confidence intervals for variable importance in random forest regression, classification, and survival. Statistics in Medicine 38, 558–582 (2019).

82. Ishwaran, H., Kogalur, U. B., Chen, X. & Minn, A. J. Random survival forests for high-dimensional data. Statistical Analysis 4, 115–132 (2011).

83. Austin, P. C. An Introduction to Propensity Score Methods for Reducing the Effects of Confounding in Observational Studies. Multivariate Behavioral Research 46, 399–424 (2011).

84. Julkunen, H. et al. Atlas of plasma NMR biomarkers for health and disease in 118,461 individuals from the UK Biobank. Nat Commun 14, 604 (2023).

