## Supplementary Methods, Tables and Figures for "Food-level dietary factors reveal cardiometabolic risk hidden by diet quality scores"

Kittrell et al.

### SUPPLEMENTARY INFORMATION

### Supplementary methods

#### 1.1 Dietary intake data processing

This section describes the construction of the person-level dietary intake matrix used as input for non-negative matrix factorization (NMF) from raw 24-hour recall data captured by the Oxford WebQ. A summary of the preprocessing pipeline is provided in the main text. The subsections below provide the full details of each processing step.

##### 1.1.1 Raw data extraction and validity filtering

Raw dietary recall data for the analytic cohort were extracted from the UK Biobank (UKB) data fields and restructured into a long-format dataset of participant x recall instance x food item x reported value. Only recall instances that passed validity checks were retained. A valid recall was defined as: intake reported as “typical” (UKB field 100020), minimum duration of five minutes (UKB field 20082), completion date available (UKB field 105010) and valid total energy intake (UKB field 26002), defined as ≥500 but ≤3,500 for females and ≥800 but ≤4,200 for males[1,2]. The cohort was restricted to participants with at least two valid recalls. The long-format dataset was then reconciled against the analytic cohort to confirm that recall counts matched.

##### 1.1.2 Variable classification and exclusion

Food and beverage variables from the Oxford WebQ were classified using UKB field metadata into ordinal variables (intake quantities) and categorical variables (food type qualifiers). The following categories of variables were excluded from the dietary matrix before further processing:

- **Alcoholic beverages** (beer/cider, red wine, rose wine, white win, fortified wine, spirits, other alcoholic drinks). Alcohol was excluded from the dietary intake matrix because it cannot simultaneously serve as a constitutive intake factor component and an adjusted confounder. Instead, mean alcohol intake (continuous grams/day, UKB field 26030) was included as a covariate in all outcome models.

- **Plain drinking water.** Water carries no caloric or nutritional information relevant to intake factor characterization and loaded indiscriminately across all factors in preliminary NMF runs.
- **Non-dietary behavioral variables.** Whether salt was added to food (UKB field 104660), and whether no fat was used for cooking (103980) were both removed as they represent preparation methods rather than nutritional content.
- **Bread butter/margarine count variables** (number of bread items with butter or margarine). These count fields record preparation method rather than independent food intake and are collinear with bread intake variables.
- **Spread type, cooking fat type and spreads/sauces.** These categorical modifier variables do not carry independent quantitative intake information in the Oxford WebQ structure.
- **Decaffeinated vs. caffeinated tea/coffee distinctions**. These sub-classifications do not contribute to nutritional differentiation.
- **Dried fruit added to cereal**. This cereal add-on field was excluded to avoid double-counting with the separate dried fruit intake variable.

##### 1.1.3 Ordinal response conversion.

Ordinal intake responses were converted to numeric servings per day. The Oxford WebQ uses several response formats: integer counts, “half” (coded as 0.5), “quarter” (coded as 0.25) and values with a “+” suffix (e.g., “6+”, treated as that integer). Responses of “Prefer not to answer”, “Varied”, “No” and standard missing indicators were treated missing. Where a participant – instance pair had no reported value for a given food item (e.g., the item was not included in the participant’s recall), intake was coded as zero (structural zero), reflecting the assumption that unreported items were not consumed during that recall period.

1.1.4 Derived food variables.

Several food items in the Oxford WebQ are recorded as base quantity paired with one or more categorical qualifiers (e.g. slices of bread x flour type). These were reconstituted into nutritionally meaningful derived variables as follows.

*Bread x flour type.* Bread intake is reported separately for four bread formats (sliced bread, baguettes, baps/pita, bread rolls), each with an associated multi-select flour-type qualifier (white, wholemeal, mixed/seeded, other). Each format’s reported portion was divided equally across the flour types selected for that format within each recall instance, producing variables such as sliced_bread_white, baguette_wholemeal, etc. These format × flour-type variables were subsequently aggregated into four bread categories (white, wholemeal, mixed/brown/seeded, other) by summing across all bread formats (see Section 1.1.5).

*Yogurt x fat type.* Total yogurt intake was split by the fat-type qualifier (low-fat, full-fat) using the same equal-allocation rule, producing yogurt_lowfat and yogurt_fullfat.

*Porridge x cooking liquid.* Porridge intake was split by the reported cooking liquid (milk, water), producing porridge_milk and porridge_water. This classification captures the cooking medium, which is distinct from milk added to cereal (captured separately in the milk reconstitution; see below).

*Potato x butter/margarine.* Boiled or baked potato intake was reclassified as potatoes_butter_marg when the participant selected the butter/margarine add-on for that recall instance; otherwise the intake was retained as potatoes.

*Milk reconstitution.* Milk is not recorded as a single intake quantity in the Oxford WebQ but must be reconstituted from multiple sources[3,4]. Total milk volume (mL) per recall instance was calculated as the sum of:

i. glasses of milk reported directly (× 250 mL per glass)

ii. milk added to cereal (× 100 mL per bowl), conditional on the cereal milk-add response (full credit for “yes”, half credit for “varied”, zero otherwise), applied across all cereal formats (porridge, muesli, oat crunch, sweet cereal, other cereal, plain cereal, bran cereal, Weetabix-style cereal)
iii. milk added to tea (× 35 mL per cup), conditional on the tea milk-add response, for standard tea and rooibos tea only (green tea, herbal tea and other tea were assumed to be consumed without milk)
iv. milk added to coffee (× 25 mL per cup), where cappuccino and latte were classified as always containing milk, while for other coffee formats (instant, filtered, espresso, other) milk was credited conditionally on the coffee milk-add response.

Total mL was converted to servings (1 serving = 250 mL = 1 glass) and allocated to the single milk type reported by the participant via the type_milk_consumed field, a single-select variable. The five milk-type categories were: whole milk, semi-skimmed milk, low-fat milk (including skimmed, powdered, and cholesterol-lowering milk), rice/oat/vegetable milk, and soya milk (with and without calcium). Participants reporting “did not have milk” received zero servings across all categories. Participants reporting “don’t know type” or “other type of milk” had their total milk volume discarded (all five type categories set to zero).

*Soup ingredient classification.* Soup servings were classified by protein source using the Oxford WebQ ingredient tag fields. For canned and homemade soups, the ingredient list was examined and classified using a hierarchical rule: meat > fish > vegetable (i.e., a soup tagged with both meat and fish was classified as meat soup). Powdered/instant soup, for which no ingredient tag is available, was classified as vegetable soup by default. This produced three soup variables (soup_meat, soup_fish, soup_veg), all of which map to a single “soups” group in the Piernas food grouping system.

*Gluten-free reallocation.* For participants self-reporting a gluten-free diet (via the special diet field, UKB field 20086), reported intake of bread items (sliced bread, baguette, bap/pitta, bread roll, garlic bread, naan bread, other bread), crackers/crispbreads and oatcakes, and white pasta was summed per recall instance into gluten-free equivalents (bread_gf, crackers_gf, pasta_gf). The corresponding regular columns for these participants were then set to zero to prevent double-counting. The gluten-free bread variable was assigned to the "bread other" aggregation group; gluten-free crackers to "snacks crackers"; and gluten-free pasta to "grains white."

1.1.5 Variable aggregation.

After deriving the complex food variables above, correlated sub-items were aggregated by summation into broader food categories to reduce redundancy and collinearity in the input matrix. The following aggregations were applied:

- **Coffee**: instant coffee + filtered coffee + espresso + other coffee + latte + cappuccino
- **Tea**: standard tea + green tea + other tea + rooibos tea + herbal tea
- **Added sweetener**: artificial sweetener added to coffee + tea + cereal
- **Added sugar**: sugar added to coffee + tea + cereal
- **Bread (mixed/brown/seeded)**: sum across all bread formats of mixed and seeded flour types
- **Bread (other)**: garlic bread + naan bread + other bread + "other" flour types across all formats + bread (gluten-free)
- **Bread (white)**: sum across all bread formats of white flour type
- **Bread (wholemeal)**: sum across all bread formats of wholemeal flour type

1.1.6 Food group mapping.

Individual food variables were mapped to food groups adapted from the classification system of Piernas et al.[4]. This system organizes Oxford WebQ food items into nutritionally coherent food groups. Our adaptation excluded alcohol (handled as a covariate), water, bread spreads,

spreads/sauces, and cooking-fat type groups, and incorporated the derived milk, soup, yogurt, porridge, potato, bread, and gluten-free variables described above. NMF was run on both the individual food level and the food group level. The complete mapping of individual foods to Piernas food groups is provided in Supplementary Table 12.

##### 1.1.7 Preprocessing pipeline for NMF.

The following sequential transformations were applied to the instance-level dietary intake matrix to produce the final NMF-ready input:

1. **Averaging across recall instances.** For each participant, intake values were averaged across all valid recall instances to produce a single person-level estimate of usual intake per food item.
2. **Coarse prevalence filter.** Food items consumed by fewer than 5% of participants (in the person-level averaged dataset) were removed. This threshold was selected as the primary analysis, with 1% and 10% thresholds tested as well.
3. **Zero-variance column removal.** Columns with zero variance in the coarse-filtered dataset were removed.
4. **Upper tail winsorization.** Values exceeding the 99$^{th}$ percentile for each variable were capped at the 99$^{th}$ percentile. Only the upper tail was winsorized; the lower tail (structural zeros from non-consumers) was left untouched.
5. **Log transformation.** Log(1 + *x*) was applied to all intake values.
6. **Standard deviation scaling.** Each variable was divided by its marginal standard deviation (SD), placing all foods on a comparable scale and preventing high-variance foods (beverages) from dominating the factorization.

When operating between the split discovery, replication and validation datasets, the transform pipeline (steps 1–6) was run on discovery participants only. The learned parameters (coarse-filter column list, 99$^{th}$ percentile cut points and marginal SDs) were saved and applied identically to the replication and validation datasets. This prevents held-out information from leaking into the NMF input matrix.

1.1.8 Quality control and validation.

Automated quality control of the dietary data included: (1) a per-variable QC report documenting raw response distributions, numeric conversion rates, missingness and structural zeros (food items not included in a given recall, treated as zero intake); (2) descriptive statistics computed at four stages of the preprocessing pipeline (pre-winsorization, post-winsorization, post-log-transformation, post-SD-scaling), including normalized Shannon entropy and top-value share to identify low-information columns; (3) a low-entropy diagnostic screen flagging columns with ≤12 unique values, normalized entropy <0.05, or >98% of values on a single level; and (4) worked validation examples from real participant records for each derived-variable rule (milk reconstitution, bread flour-type splitting, yogurt fat-type splitting, porridge liquid-type splitting, potato butter/margarine classification, soup ingredient classification, gluten-free reallocation), allowing every derivation rule to be verified by eye against actual reported values.

1.2 Variable coding

The following variable recoding was carried out before multiple imputation. In contrast to variables with genuine missingness, responses of “Prefer not to answer” (PNA) were treated as informative non-responses and retained as explicit factor levels for categorical covariates (annual household income, smoking status, ethnicity, highest educational qualification and supplement use) rather than recoded as missing. This ensures that PNA responses are not replaced with imputed values. Responses of “Do not know” and “Not applicable” were recoded as NA and multiply imputed. All nominal categorical variables were coded as unordered factors. *Assessment center instance selection*. For covariates collected at multiple assessment center visits (instances 0–3), the value from the assessment center instance closest to, and on or before, each participant’s baseline date (date of last valid dietary recall) was selected. This ensures that covariate values reflect the participant’s status at the time of dietary assessment rather than a temporally remote visit. Where no pre-baseline assessment center instance was available, the covariate value was set to missing for imputation.

*Ethnicity.* Responses to the baseline UK Biobank self-reported ethnicity question (field ID 21000) were condensed into five categories (Black, Asian, White, Mixed, Other), largely following previous research in the UK Biobank[REF 1]. This condensation further collapses all reported Asian ethnic backgrounds into a single response category. Response categories were re-coded as follows: Black = “Black or Black British”, “Caribbean”, “African”, “Any other Black background”; White = “White”, “British”, “Irish”, “Any other white background”; Mixed = “Mixed”, “White and Black Caribbean”, “White and Black African”, “White and Asian”, “Any other mixed background”; Asian = “Asian or Asian British”, “Chinese”, “Indian”, “Pakistani”, “Bangladeshi”, “Any other Asian background”; Other = “Other ethnic group.” For the primary analysis, this five-category classification was further collapsed into a binary White vs. Non-white variable. PNA responses were retained as a separate level; “Do not know” was set to missing for imputation.

*Annual household income.* Income was coded as a five-level ordinal variable (<£18,000; £18,000–30,999; £31,000–51,999; £52,000–100,000; >£100,000). Both “Do not know” and PNA responses were collapsed into a single “Prefer not to answer” factor level.

*Smoking status.* Smoking was coded as never, previous, or current. PNA was retained as an explicit factor level. "Do not know" was set to missing for imputation.

*Supplement use.* Vitamin and mineral supplement use (field ID 6155) was derived from a mark-all-that-apply field. Any specific supplement ticked was coded as “Yes”; a response of “None of the above” was coded as “No”; PNA was retained as a separate factor level. When all responses were non-response values other than PNA, supplement use was set to missing for imputation.

*Body mass index.* BMI ($kg/m^2$) was taken from the assessment center instance closest to baseline. Values outside the plausible range of 12–70 $kg/m^2$ were set to missing for imputation.

*Mean energy intake.* Total energy intake (kcal/day, converted from kJ) was computed as the mean across all valid recall instances. Implausible values were flagged using sex-specific Willett bounds (men: <800 or >4,200 kcal/day; women: <500 or >3,500 kcal/day) and retained in the dataset with a flag variable.

*Alcohol intake.* Alcohol intake was derived as the mean alcohol consumption (g/day) across valid 24-hour recalls from the Oxford WebQ nutrient data. This continuous variable was included as a covariate in outcome models rather than as part of the dietary pattern matrix (see Section 1.1). Negative values were set to missing for imputation.

*Family history.* Family history of heart disease and diabetes was derived from parental illness fields (father and mother illness). Each was coded as binary (1 = at least one parent with the condition, 0 = no parental history). Participants for whom all parental illness responses were non-response values were set to missing for imputation. Both family history variables were included as covariates for all primary outcomes.

*Prevalent hypertension and dyslipidemia.* Baseline hypertension and dyslipidemia flags were composite indicators combining ICD-10 codes, self-reported conditions, and relevant medication use, constructed in the phenotyping pipeline. These variables are complete by design (default = no disease) and were not imputed.

#### 1.3 Derived variables

Several derived variables were calculated in our UKB dataset. Variables derived from raw sub-fields were computed post-imputation to preserve uncertainty; the raw sub-fields were carried forward as ordered or unordered factors for imputation, and the original sub-fields used to construct each derived variable were then excluded from the analytic dataset.

*Education.* Following previous research in the UKB[3,5], an education years variable was created by converting the responses from the education qualifications variable (field ID 6138) into the equivalent years of education. Response categories were mapped onto years of education using the International Standard Classification of Education (ISCED) scale as follows: 7 years = "none of the above (no qualifications)"; 10 years = "CSEs or equivalent" or "O levels/GCSEs or equivalent"; 13 years = "A levels/AS levels or equivalent"; 15 years = "other professional qualification"; 19 years = "NVQ or HND or HNC or equivalent"; 20 years = "college or university degree." Where participants marked multiple educational qualifications, the qualification with the highest corresponding years of education was used for that participant. Raw qualification dummies were carried forward as binary variables for imputation, and the education years derivation was applied within each imputed dataset. Participants with PNA responses were assigned to a "Prefer not to answer" factor level for the categorical education variable and excluded from the years-of-education derivation.

*Leisure time physical activity.* As has been done previously[3,6], we used responses from a mark-all-that-apply question (UKB field 6164) asking participants about types of physical activity they have undertaken in the past 4 weeks to create a summary leisure time physical activity (LTPA) score. For each type of activity, if the participant reported undertaking that activity in the past 4 weeks then they were asked how many times in the past 4 weeks and how long they spent on the activity each time. Those responses were used to determine the number of days/week for each activity and the number of minutes for each activity. These values were multiplied and divided by 7 to get the mins/day for each activity, which was multiplied by the metabolic equivalent of task (MET) for each activity: walking = 3.3 MET, light DIY = 2.25 MET, heavy DIY = 4.5 MET, sports/exercise = 8.0 MET, other exercise = 4.5 MET. We then summed these values across all activities to get a total LTPA MET value for each participant. Participants were categorized into three groups of LTPA using the IPAQ scoring system: (1) high activity: ≥3,000 MET-mins per week; (2) moderate: 600–2,999 MET-mins per week; and (3) low: <600 MET-mins per week. Raw frequency and duration responses for each activity domain were carried forward as ordered factors for imputation (using proportional odds logistic regression), and the LTPA derivation was applied within each imputed dataset.

*Energy intake quintile.* Quintile boundaries for mean energy intake were computed from the observed (pre-imputation) distribution. These boundaries were then applied within each imputed dataset to create an energy quintile variable used as a covariate in the multivariable adjusted Cox models.

1.4 Variable quality control.

As a quality control step conducted after imputation but before running Cox proportional hazards models, we systematically examined the crosstabs between each categorical covariate and the event indicator for each primary outcome (CAD, T2D, CKD) among at-risk participants. Any categorical variable with fewer than 5 events for a single response level was flagged for further inspection, as sparse cells can compromise model stability and produce unreliable effect estimates. This check was performed independently within each of the five imputed datasets. Additionally, columns with ≥80% missing data were excluded from the imputation model, with the exception of physical activity sub-fields, which were retained because they are the constituent inputs to the LTPA derivation and cannot be meaningfully replaced by a single summary measure.

### Supplementary figures

a

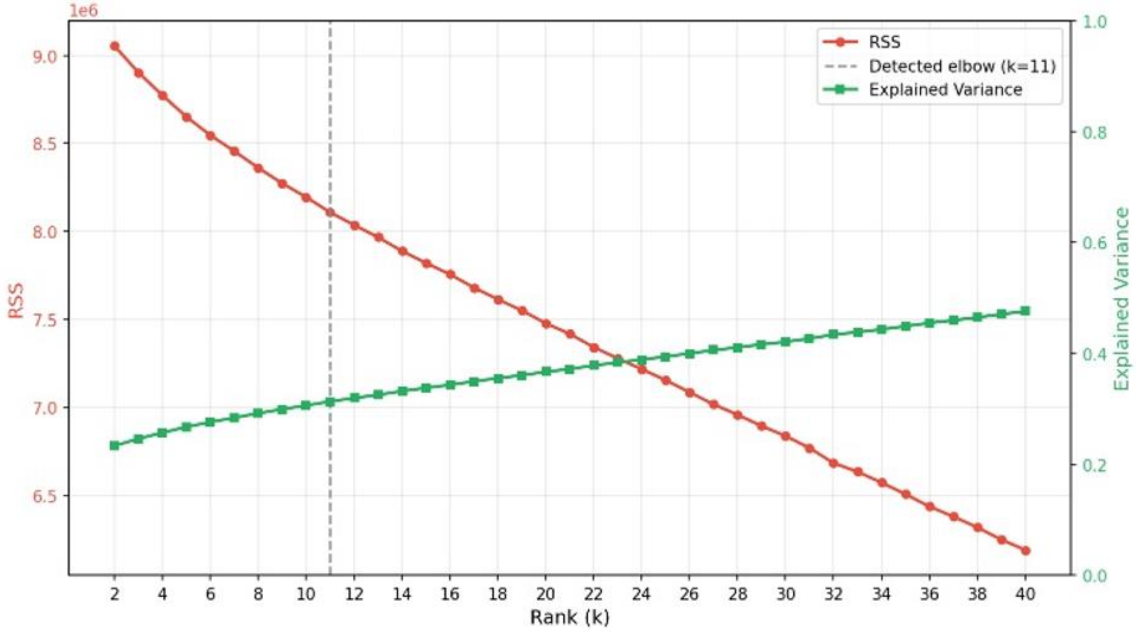

b

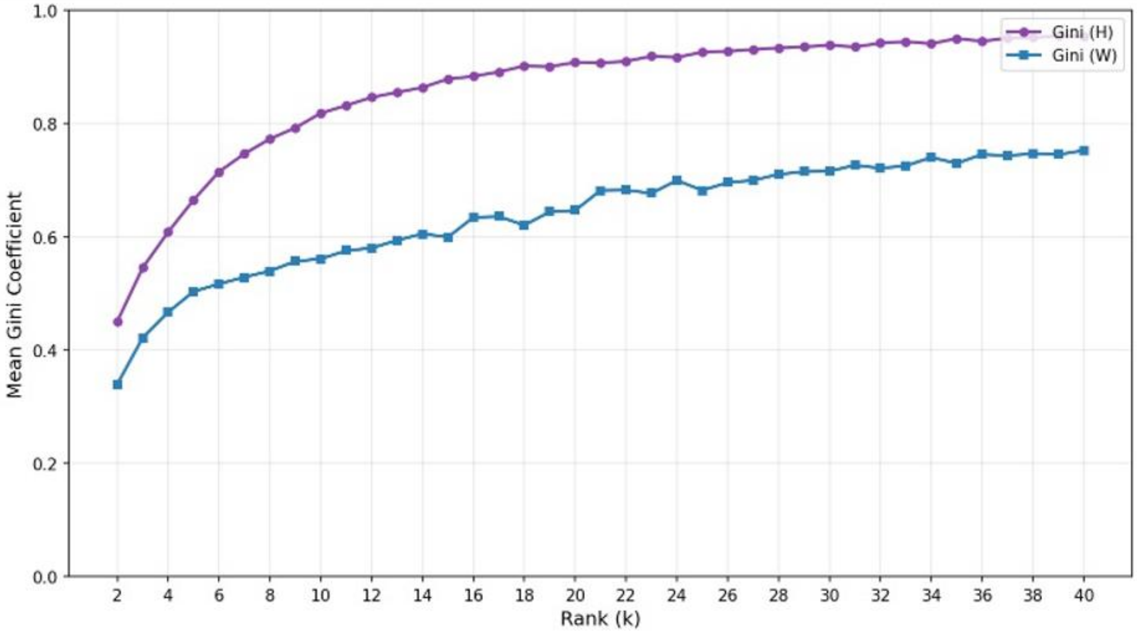

**Supplementary Figure 1. Rank selection diagnostics.** (a) Explained variance (proportion of total Frobenius norm reconstructed) and (b) Gini sparsity index of the H-matrix as a function of candidate rank *k* = 2–40, computed using deterministic NNDSVD initialization without regularization on the discovery dataset (n = 55,839). The elbow region used to identify the candidate rank range for stability testing is indicated.

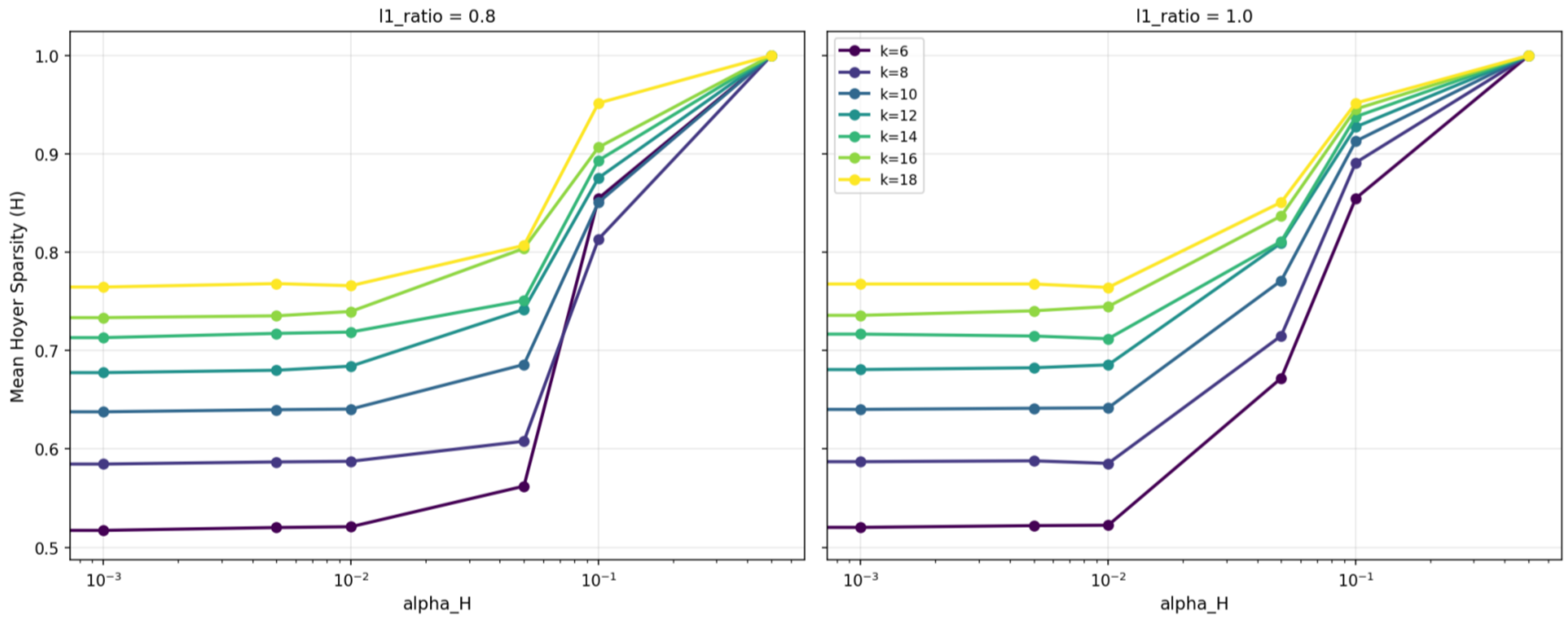

**Supplementary Figure 2. L1 regularization sensitivity**. Mean Hoyer sparsity of the H-matrix as a function of the regularization strength (α_H) for candidate ranks k = 6–18 at l1_ratio = 0.8 (left) and l1_ratio = 1.0 (right). Sparsity remains stable at low regularization strengths and increases sharply above α_H ≈ $10^{-2}$, confirming that the unregularized factorization already produces a naturally sparse solution.

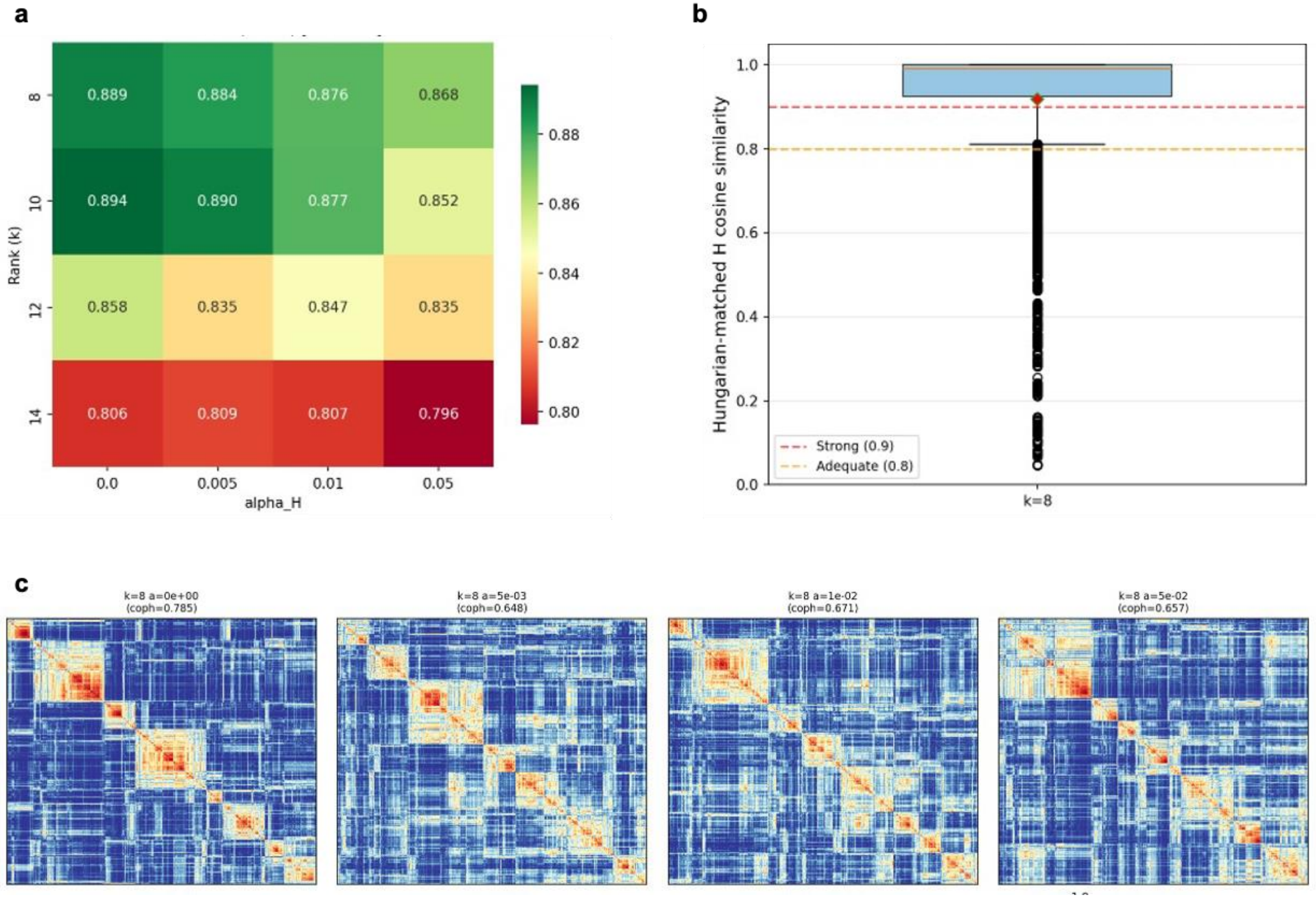

**Supplementary Figure 3. Factor stability assessment at *k* = 8.** (a) Pairwise cosine similarity of H-matrix rows across 30 random initializations at *k* = 8, showing convergence to a stable factor structure. (b) Split-half stability: cosine similarity between factors derived from 10 independent random 50/50 splits of the discovery dataset. (c) Consensus heatmap showing the frequency within which each pair of foods co-loaded on the same factor across 30 runs. Coph; cophenetic correlation.

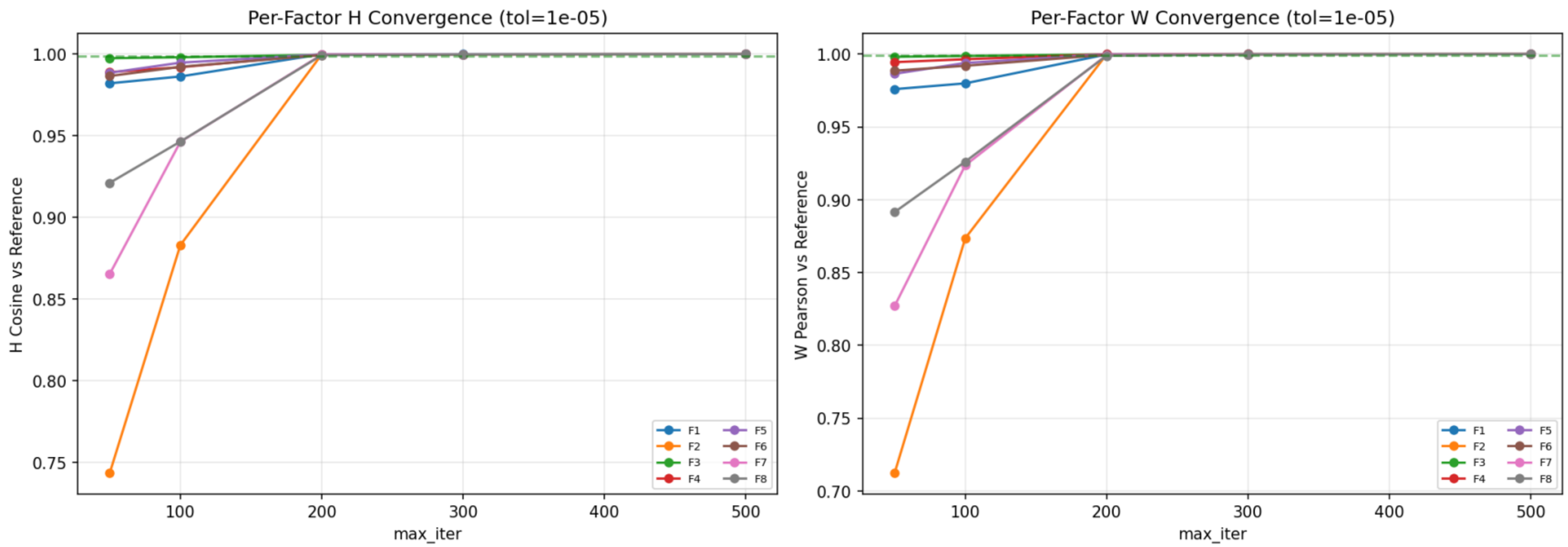

**Supplementary Figure 4. NMF convergence diagnostics.** Per-factor convergence as a function of maximum iterations (tol = $1 \times 10^{-5}$) for the H-matrix (left, cosine similarity vs. reference) and W-matrix (right, Pearson correlation vs. reference) during the rank selection process. All eight factors converge to cosine similarity and correlation > 0.99 by 500 iterations, with most factors stabilizing by 200 iterations.

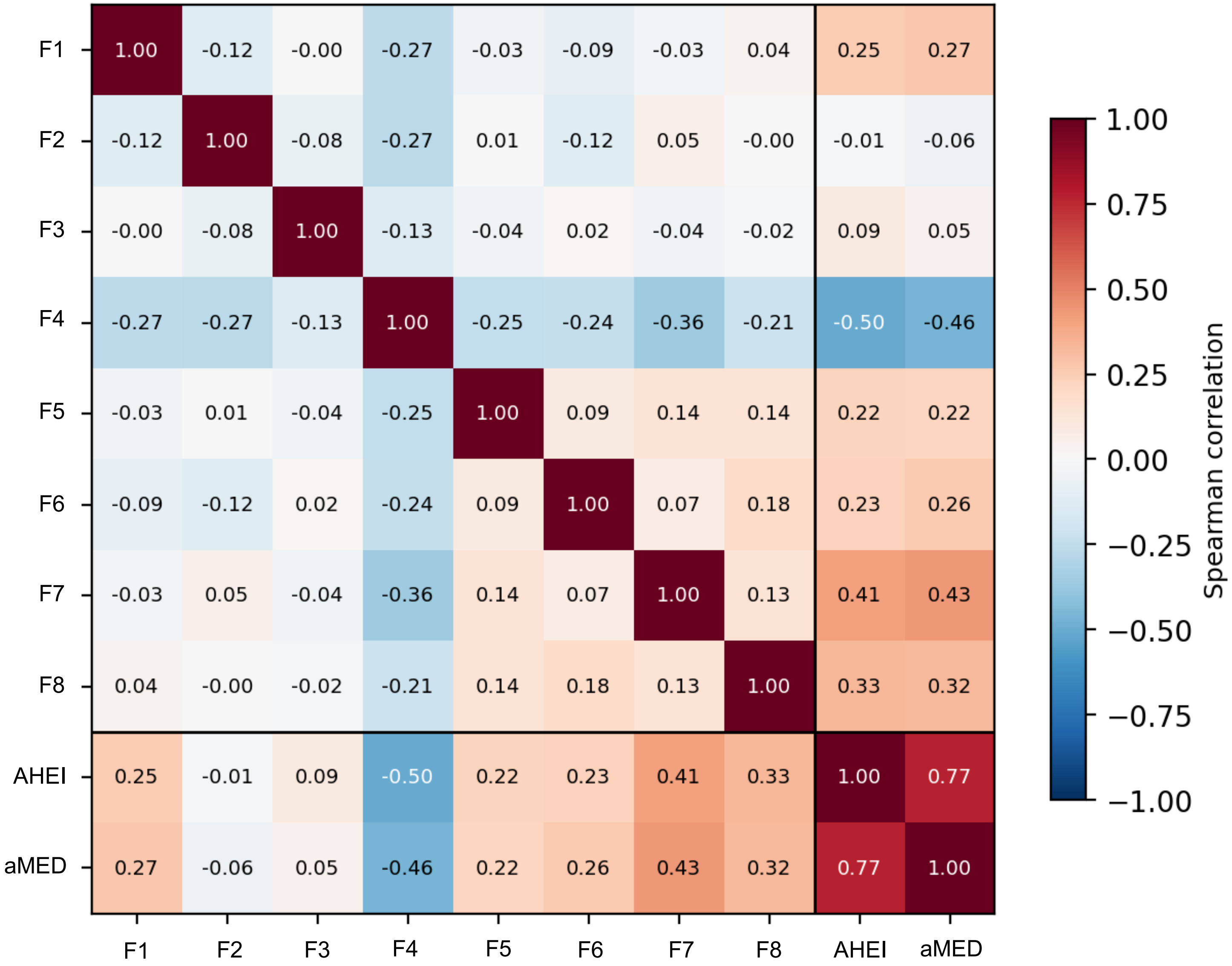

**Supplementary Figure 5. Correlations among NMF factors and diet quality scores.** Pairwise Spearman correlations among the eight NMF factor loadings and energy-adjusted AHEI-2010 and aMED scores in the discovery dataset (n = 55,839).

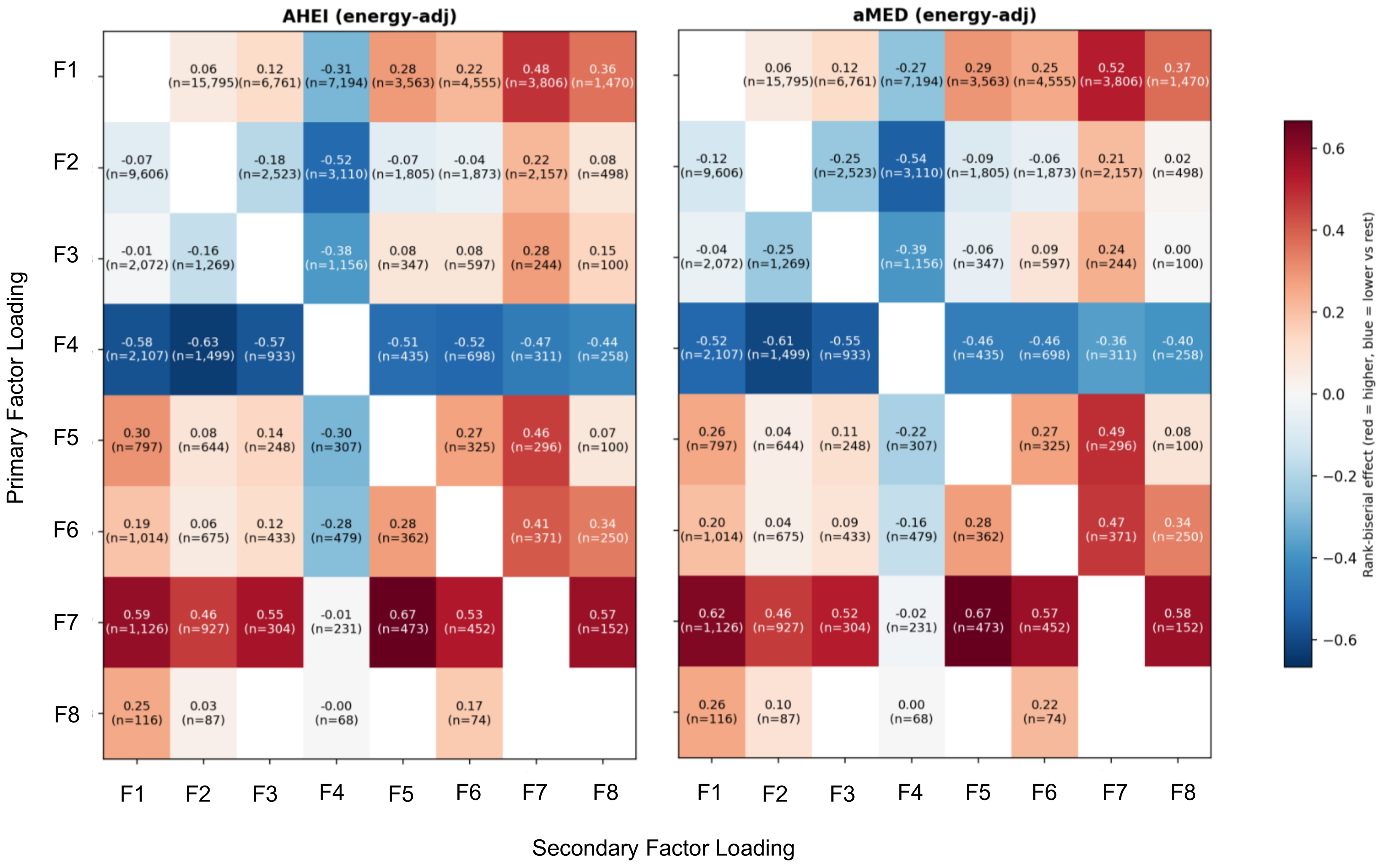

**Supplementary Figure 6. Diet quality scores by primary and secondary factor combination.** Heatmap showing mean energy-adjusted (**left**) AHEI-2010 and (**right**) aMED scores for each combination of dominant (primary) and secondary NMF factor loading. Cells with fewer than 30 individuals are masked.

### Supplementary tables

**Supplementary Table 1.** Baseline descriptive statistics – UK Biobank participants included in versus excluded from the analytic cohort.

| | **Diet-Valid Cohort (N = 102,316)** | **Analytical Cohort (N = 87,162)** |
|---|---|---|
| Age at baseline | 59.8 (7.8) | 59.2 (7.8) |
| Female | 56,730 (55.4%) | 49,719 (57.0%) |
| **Ethnicity** | | |
| White | 99,013 (97.1%) | 84,439 (97.2%) |
| Asian | 1,173 (1.2%) | 925 (1.1%) |
| Black | 733 (0.7%) | 609 (0.7%) |
| Mixed | 502 (0.5%) | 444 (0.5%) |
| Other | 567 (0.6%) | 473 (0.5%) |
| Income >100k | 6,806 (6.7%) | 6,194 (7.1%) |
| Education, years | 16.5 (4.5) | 16.6 (4.4) |
| Family history of diabetes | 17,682 (17.3%) | 14,772 (16.9%) |
| Family history of CVD | 44,952 (43.9%) | 38,026 (43.6%) |
| Prevalent hypertension | 52,491 (51.3%) | 41,623 (47.8%) |
| Prevalent dyslipidemia | 53,311 (52.1%) | 42,379 (48.6%) |
| Supplement use | 33,362 (32.6%) | 28,560 (32.8%) |
| BMI, kg/m$^2$ | 26.6 (4.6) | 26.3 (4.3) |
| Never smoker | 58,698 (57.4%) | 51,373 (58.9%) |
| Physical activity, high | --- | 29,408 (33.7%) |
| Alcohol intake, g/day | 15.1 (18.6) | 15.1 (18.5) |
| Energy intake, kcal/day | 2,027 (471) | 2,027 (471) |
| Protein, g/day | 80.1 (19.5) | 79.9 (19.4) |
| Fat, g/day | 71.7 (23.5) | 71.7 (23.6) |
| Carbohydrate, g/day | 251.9 (65.8) | 252.0 (65.8) |
| Fiber, g/day | 18.3 (5.8) | 18.3 (5.8) |
| Saturated Fat, g/day | 26.4 (10.1) | 26.5 (10.2) |
| AHEI score, energy-adjusted | 48.6 (12.7) | 48.6 (12.7) |
| aMED score, energy -adjusted | 4.0 (1.8) | 4.0 (1.8) |

**Supplementary Table 2.** Baseline descriptive statistics of UK Biobank participants split by discovery, replication and validation datasets.

| | **Discovery (n = 55,839)** | **Replication (n = 23,932)** | **Validation (n = 7,391)** |
|---|---|---|---|
| Age at baseline | 59.2 (7.8) | 59.2 (7.8) | 59.5 (7.7) |
| Female | 31,872 (57.1%) | 13,660 (57.1%) | 4,187 (56.6%) |
| **Ethnicity** | | | |
| White | 53,967 (97.0%) | 23,165 (97.1%) | 7,307 (99.1%) |
| Asian | 622 (1.1%) | 283 (1.2%) | 20 (0.3%) |
| Black | 423 (0.8%) | 177 (0.7%) | 9 (0.1%) |
| Mixed | 320 (0.6%) | 111 (0.5%) | 13 (0.2%) |
| Other | 330 (0.6%) | 122 (0.5%) | 21 (0.3%) |
| Income >100k | 3,953 (7.0%) | 1,757 (7.3%) | 502 (6.8%) |
| Education, years | 16.5 (4.4) | 16.5 (4.4) | 17.1 (4.2) |
| Family history of diabetes | 9,434 (16.9%) | 4,036 (16.9%) | 1,072 (14.5%) |
| Family history of CVD | 23,878 (42.8%) | 10,302 (43.0%) | 3,282 (44.4%) |
| Prevalent hypertension | 27,074 (48.5%) | 11,562 (48.3%) | 2,987 (40.4%) |
| Prevalent dyslipidemia | 27,146 (48.6%) | 11,687 (48.8%) | 3,546 (48.0%) |
| Supplement use | 18,220 (32.6%) | 7,917 (33.1%) | 2,420 (32.7%) |
| BMI, kg/$m^2$ | 26.3 (4.3) | 26.3 (4.3) | 26.3 (4.3) |
| Never smoker | 32,661 (58.5%) | 14,150 (59.1%) | 4,552 (61.6%) |
| Physical activity, high | 4,826 (8.6%) | 2,037 (8.5%) | 535 (7.2%) |
| Alcohol intake, g/day | 15.2 (18.5) | 15.3 (18.7) | 14.2 (17.6) |
| Energy intake, kcal/day | 2,026 (472) | 2,025 (471) | 2,044 (469) |
| Protein, g/day | 79.9 (19.5) | 79.9 (19.4) | 79.7 (19.5) |
| Fat, g/day | 71.6 (23.6) | 71.6 (23.6) | 72.8 (23.8) |
| Carbohydrate, g/day | 251.7 (65.8) | 251.5 (65.9) | 255.8 (65.4) |
| Fiber, g/day | 18.3 (5.8) | 18.3 (5.8) | 18.3 (5.8) |
| Saturated Fat, g/day | 26.4 (10.1) | 26.4 (10.1) | 27.1 (10.2) |
| AHEI score, energy-adjusted | 48.7 (12.6) | 48.7 (12.7) | 47.5 (12.7) |
| aMED score, energy -adjusted | 4.1 (1.8) | 4.1 (1.8) | 4.0 (1.8) |

**Supplementary Table 3.** Alternative Healthy Eating Index-2010 (AHEI) component scoring criteria and corresponding food fields in the UK Biobank.

| Component | Food Fields | Criteria for 0 points | Criteria for 10 points |
|---|---|---|---|
| Vegetables (servings/d) | Allium vegetables, Green leafy/cabbages, Raw salad, Root vegetables, Tomatoes, Other vegetables, including mushrooms, fruiting and mixed vegetables | 0 | ≥5 |
| Whole Fruit (servings/d) | Apples & pears, Berries, Citrus, Dried fruit, Other fruit, Stewed fruit | 0 | ≥4 |
| Whole grains (servings/d) | Wholemeal bread, Bran cereal, Oat cereal (non-sugar), Muesli, Wholemeal pasta, Brown rice & other grains, Biscuit cereal, Savory crackers | 0 | Women ≥5<br>Men ≥6 |
| Sugar-sweetened beverages (servings/d) | | ≥1 | 0 |
| Nuts & legumes (servings/d) | Unsalted nuts & seeds, Salted nuts & seeds, Legumes & pulses, Peas/Sweetcorn, Soy desserts and yogurt, Meat substitutes – Vegetarian alternatives | 0 | ≥1 |
| Red & processed meat (servings/d) | Beef, Lamb, Pork, Other meat, Bacon, Ham, Liver, Sausage | ≥1.5 | 0 |
| Seafood (servings/wk) | Breaded/battered Fish, Oily fish, White fish & tinned tuna, Sushi, Shellfish | 0 | ≥2 |
| Trans fat (% of energy) | -- Not available -- | --- | --- |
| PUFA (% of energy) | n-3 fatty acids, n-6 farry acids (Estimated food nutrients) | ≤2% energy | ≥10% energy |
| Sodium (mg/d) | Sodium (Estimated food nutrients) | Highest decile | Lowest decile |
| Alcohol (drinks/d) | Beer & cider, Spirits, Fortified wine, Red wine, White wine | Women ≥2.5<br>Men ≥3.5 | 0.5–1.5<br>0.5–2.0 |
| **Total** | | **0** | **100** |

Scoring criteria for the AHEI-2010 as adapted for the UK Biobank. Each component is scored continuously from 0 (least healthy) to 10 (most healthy), producing a total score ranging from 0 to 100. Scoring criteria are based on servings/d unless otherwise noted. Adaptations from the original Chiuve et al.[7] (2012) scoring are noted. PUFA, polyunsaturated fatty acids; n-3, omega-3; n-6, omega-6; mg/d, milligrams per day.

**Supplementary Table 4.** Alternate Mediterranean Diet Score (aMED) component scoring criteria and corresponding food fields in the UK Biobank.

| Component | Food Fields | Criteria for 0 points | Criteria for 1 point |
|---|---|---|---|
| Vegetables (servings/d) | Allium vegetables, Green leafy/cabbages, Raw salad, Root vegetables, Tomatoes, Other vegetables, including mushrooms, fruiting and mixed vegetables | Below the median | Above the median |
| Legumes (servings/d) | Legumes & pulses, Peas/Sweetcorn, Soy desserts and yogurt, Meat substitutes – Vegetarian alternatives | Below the median | Above the median |
| Fruit (servings/d) | Apples & pears, Berries, Citrus, Dried fruit, Other fruit, Stewed fruit | Below the median | Above the median |
| Nuts (servings/d) | Unsalted nuts & seeds, Salted nuts & seeds | Below the median | Above the median |
| Whole grains (servings/d) | Wholemeal bread, Bran cereal, Oat cereal (non-sugar), Muesli, Wholemeal pasta, Brown rice & other grains, Biscuit cereal, Savory crackers | Below the median | Above the median |
| Fish (servings/d) | Breaded/battered Fish, Oily fish, White fish & tinned tuna, Sushi, Shellfish | Below the median | Above the median |
| MUFA:SFA | Monounsaturated fatty acids, Saturated fatty acids (Estimated food nutrients) | Below the median | Above the median |
| Red & processed meat (servings/d) | Beef, Lamb, Pork, Other meat, Bacon, Ham, Liver, Sausage | Above the median | Below the median |
| Alcohol (g/d) | Alcohol consumption (Estimated food nutrients) | < 5 or > 25 | 5–25 |
| **Total** | | **0** | **9** |

Scoring criteria for the aMED as adapted for the UKB. Standard components score 1 point if intake exceeds the cohort-specific median; red and processed meat is reverse-scored (1 point if below median); alcohol uses fixed cut points (5–25 g/day). Fruit juice is not included in the Fruit component. Total score ranges from 0 (least adherent) to 9 (most adherent). Following Trichipoulou et al.[8] as adapted by Tessier et al.[9]. MUFA, monounsaturated fatty acid; SFA, saturated fatty acid; g/d, grams per day.

**Supplementary Table 5.** Data sources and diagnostic criteria for prevalent and incident disease definitions.

| **Chronic Disease** | **Baseline measures (field ID)** | **Baseline verbal interview diagnosis codes** | **ICD-10 codes** | **ICD-9 codes** |
|---|---|---|---|---|
| Colorectal cancer | - | 1022, 1023 | C18-20 | 153, 154 |
| Lung cancer | - | 1001, 1027, 1028, 1080 | C30, C34 | 162 |
| Esophageal cancer | - | 1017 | C15 | 150 |
| Liver cancer | - | 1024 | C22 | 155 |
| Pancreatic cancer | - | 1026 | C25 | 157 |
| Brain cancer | - | 1032 | C71 | 191 |
| Leukemia | - | 1048 | C91-C95 | 204-208 |
| Lymphoma | - | 1047 | C81-C86, C88 | 201-203 |
| Breast cancer | - | 1002 | C50 | 174, 175 |
| Ovarian cancer | - | 1039 | C56 | 183 |
| Prostate cancer | - | 1044 | C61 | 185 |
| Coronary artery disease | Heart attack diagnosis by physician (6150)<br>Self-report (20004) of coronary angioplasty [1070], coronary artery bypass grafts [1095], or triple heart bypass [1523] | 1075 | I21-I25 | 412, 414 |
| Type 2 diabetes | Taking insulin medication (6153, 6177)<br>Diabetes diagnosed by physician (2443)<br>Non-fasting blood HbA1c ≥ 48 mmol/mol (30750)<br>Non-fasting blood glucose ≥ 11.1 mmol/L (30740) | 1223 | E11 | - |
| Other diabetes mellitus | - | 1220, 1222 | E10, E12-E14 | - |
| Chronic kidney diseases | - | 1192, 1193, 1194 | N18 | 585 |
| **Clinical Risk Factors** | **Baseline measures (field ID)** | **Baseline verbal interview diagnosis codes** | **ICD-10 codes** | **ICD-9 codes** |
| Hypertension | High blood pressure diagnosis by physician (6150)<br>Taking medication for high blood pressure (6153, 6177)<br>Blood pressure SBP/DBP ≥ 140/90 mmHg (4079, 4080) | 1065, 1072 | I10-I13, I15 | 401-405 |
| Dyslipidemia | Taking cholesterol lowering medication (6153, 6177)<br>Blood cholesterol ≥ 240 / 38.67 (30690)<br>Blood LDL ≥ 160 / 38.67 (30780)<br>Blood triglycerides ≥ 200 / 88.57 (30870) | 1473 | - | - |

Verbal interview diagnosis codes are contained in the cancer (field ID 20001) and non-cancer illness (field ID 20002) variables. Incident disease cases were identified using ICD-10/ICD-9 codes and corresponding date of diagnosis taken from linked hospital inpatient records and from primary cause of death information from linked death register data. Baseline prevalence for all diseases and clinical risk factors was calculated for all participants using baseline measures (including verbal interview diagnosis codes) + those with an ICD diagnosis before or on the baseline date.

Incident cases are defined as those with an ICD date of diagnosis after the baseline date who do not have any prevalent diagnosis. Unless specific ICD subcategories are already given with dot separators, all ICD codes listed also include all subcategories (e.g. ). HbA1c: hemoglobin A1c; SBP: systolic blood pressure; DBP: diastolic blood pressure.

**Supplementary Table 6.** Variable missingness.

| Variable | Missing, N (%) |
|---|---|
| Leisure-time physical activity | 3,474 (3.99%) |
| Family history of diabetes | 1,355 (1.55%) |
| Family history of heart disease | 1,355 (1.55%) |
| Years of education | 261 (<1%) |
| Body mass index | 178 (<1%) |
| Household income | 101 (<1%) |
| Ethnicity | 57 (<1%) |
| Supplement use | 22 (<1%) |
| Smoking status | 21 (<1%) |
| Mean energy intake | 0 (0%) |
| Sex | 0 (0%) |
| Age at baseline | 0 (0%) |
| Mean alcohol intake | 0 (0%) |
| Nation of recruitment | 0 (0%) |
| Prevalent dyslipidemia | 0 (0%) |
| Prevalent hypertension | 0 (0%) |

**Supplementary Table 7.** Clinical biomarker summary statistics by country of recruitment.

| | **England (n = 79,771)** | **Scotland/Wales (n = 7,391)** | **Total (n = 87,162)** |
|---|---|---|---|
| Albumin (g/L) | 45.5 (2.6) | 45.3 (2.5) | 45.5 (2.6) |
| ALP (U/L) | 80.6 (23.2) | 80.5 (23.9) | 80.6 (23.3) |
| Alpha-1-antitrypsin (g/L) | 25.6 (9.0) | 25.5 (11.4) | 25.6 (9.3) |
| ALT (U/L) | 22.1 (12.7) | 22.9 (12.3) | 22.2 (12.7) |
| ApoA (g/L) | 1.57 (0.27) | 1.55 (0.26) | 1.57 (0.27) |
| ApoB (g/L) | 1.04 (0.23) | 1.04 (0.23) | 1.04 (0.23) |
| BMI (kg/m²) | 26.3 (4.3) | 26.4 (4.3) | 26.3 (4.3) |
| Body fat % (%) | 30.3 (8.4) | 30.4 (8.4) | 30.3 (8.4) |
| Calcium (mmol/L) | 2.38 (0.09) | 2.36 (0.09) | 2.38 (0.09) |
| Creatinine (µmol/L) | 71.2 (13.7) | 70.5 (13.9) | 71.1 (13.8) |
| CRP (mg/L) | 2.12 (3.72) | 2.17 (3.92) | 2.12 (3.74) |
| Cystatin C (mg/L) | 0.88 (0.13) | 0.87 (0.13) | 0.88 (0.13) |
| Diastolic BP (mmHg) | 81.6 (10.0) | 82.7 (10.1) | 81.7 (10.0) |
| Direct bilirubin (µmol/L) | 1.83 (0.80) | 1.81 (0.77) | 1.82 (0.80) |
| GGT (U/L) | 32.7 (32.3) | 33.3 (38.4) | 32.7 (32.8) |
| Glucose (mmol/L) | 4.98 (0.63) | 4.84 (0.65) | 4.97 (0.63) |
| HbA1c (mmol/mol) | 34.6 (3.6) | 34.4 (3.6) | 34.6 (3.6) |
| HDL-C (mmol/L) | 1.51 (0.38) | 1.48 (0.37) | 1.51 (0.38) |
| IGF-1 (nmol/L) | 22.0 (5.5) | 21.9 (5.6) | 22.0 (5.6) |
| LDL-C (mmol/L) | 3.61 (0.82) | 3.63 (0.82) | 3.61 (0.82) |
| Lipoprotein(a) (nmol/L) | 43.6 (48.6) | 44.5 (49.3) | 43.7 (48.6) |
| Phosphate (mmol/L) | 1.17 (0.16) | 1.13 (0.16) | 1.16 (0.16) |
| Systolic BP (mmHg) | 136 (18) | 137 (19) | 136 (18) |
| TG/HDL ratio | 1.23 (0.98) | 1.26 (1.02) | 1.23 (0.99) |
| Total bilirubin (µmol/L) | 9.34 (4.53) | 9.23 (4.28) | 9.33 (4.51) |
| Total cholesterol (mmol/L) | 5.79 (1.07) | 5.79 (1.06) | 5.79 (1.07) |

| | England (n = 79,771) | Scotland/Wales (n = 7,391) | Total (n = 87,162) |
|---|---|---|---|
| Triglycerides (mmol/L) | 1.63 (0.94) | 1.63 (0.97) | 1.63 (0.94) |
| Urate (µmol/L) | 300 (77) | 298 (78) | 300 (77) |
| Urea (mmol/L) | 5.28 (1.22) | 5.21 (1.23) | 5.27 (1.22) |
| Vitamin D (nmol/L) | 50.7 (20.7) | 43.4 (20.0) | 50.0 (20.7) |
| Waist circumference (cm) | 87.4 (12.7) | 86.5 (12.5) | 87.3 (12.7) |
| Waist-hip ratio | 0.85 (0.09) | 0.85 (0.08) | 0.85 (0.09) |
| WBC ($10^9$ cells/L) | 6.65 (1.69) | 6.56 (1.69) | 6.64 (1.69) |

Values are mean (SD). BMI, body mass index; HbA1c, glycated haemoglobin; ApoA, apolipoprotein A; ApoB, apolipoprotein B; HDL-C, high-density lipoprotein cholesterol; LDL-C, low-density lipoprotein cholesterol; TG, triglycerides; CRP, C-reactive protein; WBC, white blood cell count; ALP, alkaline phosphatase; ALT, alanine aminotransferase; GGT, gamma-glutamyltransferase; BP, blood pressure; IGF-1, insulin-like growth factor 1.

**Supplementary Table 8.** Components and calculation of LTPA scores

| LTPA (field ID) | METs |
|---|---|
| Walking for pleasure (6164, 981, 971) | 3.3 METs * mins/week |
| Strenuous sports (6164, 1001, 991) | 8.0 METs * mins/week |
| Other exercises (6164, 3647, 3637) | 4.5 METs * mins/week |
| Light DIY (6164, 1021, 1011) | 2.25 METs * mins/week |
| Heavy DIY (6164, 2634, 2624) | 4.5 METs * mins/week |

LTPA: leisure-time physical activity; MET: metabolic equivalent of task; DIY: do-it-yourself. METs used for each type of activity were taken from Argentieri et al. (2025).

**Supplementary Table 9.** Interpretation of NMF-derived latent intake factors.

| Intake Factor | % | Non-zero & inactive (%) | Major Food | % | Major Food | % |
|---|---|---|---|---|---|---|
| Factor 1 *Prudent base* | 27.8 | 40% | Tea | 8.2 | Porridge with milk | 3.2 |
| | | | Bananas | 6.6 | Satsuma | 2.7 |
| | | | Apples | 5.8 | Pears | 2.6 |
| | | | Wholemeal bread | 5.5 | Whole wheat cereal | 2.5 |
| | | | Low-fat yogurt | 3.7 | Potatoes | 2.4 |
| | | | Oranges | 3.3 | | |
| Factor 2 *Cheese & whole grains* | 9.7 | 51% | Coffee | 6.3 | Orange juice | 3.2 |
| | | | Hard cheese | 4.8 | Blue cheese | 3.2 |
| | | | Mixed, brown, seeded bread | 4.7 | Stewed fruit | 2.9 |
| | | | Muesli | 4.3 | Crackers | 2.6 |
| | | | Soft cheese | 3.5 | Cake | 2.5 |
| Factor 3 *Meat & starchy vegetables* | 2.5 | 39% | Carrots | 8.0 | Yorkshire pudding | 3.9 |
| | | | Broccoli | 5.8 | Potatoes | 3.6 |
| | | | Cauliflower | 5.8 | Green beans | 3.4 |
| | | | Cabbage/kale | 5.0 | Pork | 3.0 |
| | | | Parsnip | 4.9 | Fried potatoes | 2.8 |
| | | | Turnips/swede | 4.2 | Peas | 2.8 |
| | | | Sprouts | 4.1 | Leeks | 2.6 |
| | | | Mashed potatoes | 3.9 | | |
| Factor 4 *High sugar & processed* | 2.3 | 55% | White bread | 4.8 | Chocolate bar | 2.8 |
| | | | Crisps | 3.8 | Fizzy drink | 2,6 |
| | | | Added sugar | 3.2 | Coffee | 2.5 |
| | | | Fried potatoes | 3.0 | Chocolate covered biscuits | 2.5 |
| | | | Tea | 2.9 | | |
| Factor 5 *Aromatic vegetables & rice* | 0.5 | 32% | Onions | 9.5 | White rice | 3.9 |
| | | | Garlic | 8.6 | Vegetable pieces | 3.4 |
| | | | Tinned tomato | 8.2 | Other vegetables | 3.0 |
| | | | Mushrooms | 6.9 | White pasta | 2.8 |
| | | | Sweet peppers | 6.9 | Leeks | 2.6 |
| | | | Zucchini | 5.3 | Butternut squash | 2.5 |
| | | | Other beans | 5.2 | | 2.8 |
| Factor 6 *Fruit & low-fat dairy* | 1.4 | 45% | Grapes | 5.1 | Plums | 3.7 |
| | | | Berries | 4.8 | Mixed fruit | 3.3 |
| | | | Low-fat yogurt | 4.6 | Low calorie drinks | 3.2 |
| | | | Peaches | 4.5 | Cherries | 3.1 |
| | | | Melons | 4.4 | Other fruit | 3.0 |
| | | | Pineapple | 4.3 | Mango | 2.9 |
| | | | Coffee | 4.2 | Bananas | 2.6 |
| Factor 7 *Plant-based & fish* | 1.0 | 55% | Unsalted nuts | 6.5 | Tea | 2.8 |
| | | | Seeds | 5.8 | Pure fruit & vegetable juice | 2.7 |
| | | | Dried fruit | 4.6 | Oatcakes | 2.6 |
| | | | Avocado | 3.8 | Oily fish | 2.6 |
| | | | Olives | 3.5 | Berries | 2.5 |
| | | | Dark chocolate | 3.3 | Other vegetables | 2.5 |

| | | | | | | |
|---|---|---|---|---|---|---|
| | | | Spinach | 3.1 | | |
| Factor 8<br>*Raw salad* | 1.1 | 28% | Lettuce | 12.4 | Coleslaw | 4.8 |
| | | | Cucumbers | 11.0 | Watercress | 4.7 |
| | | | Fresh tomato | 10.0 | Sweet peppers | 4.6 |
| | | | Side salad | 9.8 | Onions | 2.6 |
| | | | Beetroot | 7.2 | Ham | 2.6 |
| | | | Celery | 6.0 | | |

Intake factor % is percent of population with extreme archetype for that factor. Non-zero and inactive are the percent of foods per factor that are non-zero but below the 2.5% relative weight active cutoff. Foods contributing ≥2.5% to the construction of each intake factor (name and % of contribution) are listed.

**Supplementary Table 10.** Generalization of NMF factor structure to held-out datasets.

| **Overall reconstruction** | | | |
|---|---|---|---|
| | **Discovery** | **Replication** | **Validation** |
| *n* | 55,839 | 23,932 | 7,391 |
| Explained variance | 0.292 | 0.292 | 0.283 |
| EV change vs. training (%) | — | −0.02% | −3.17% |
| Relative reconstruction error | 0.842 | 0.842 | 0.847 |
| Mean loading correlation (r) | — | >0.999 | 0.993 |
| **Per-factor loading distributions (Kolmogorov–Smirnov tests)** | | | |
| **Factor** | **KS statistic** (replication) | **KS statistic** (validation) | *P* (validation) |
| F1 (Prudent base) | 0.004 | 0.010 | 0.566 |
| F2 (Cheese & whole grains) | 0.008 | 0.041 | <0.001 |
| F3 (Meat & starchy vegetables) | 0.005 | 0.079 | <0.001 |
| F4 (High sugar & processed foods) | 0.005 | 0.014 | 0.182 |
| F5 (Aromatic vegetables & rice) | 0.005 | 0.038 | <0.001 |
| F6 (Fruit & low-fat dairy) | 0.005 | 0.040 | <0.001 |
| F7 (Plant-based & fish) | 0.006 | 0.015 | 0.123 |
| F8 (Raw salad) | 0.006 | 0.039 | <0.001 |

The H-matrix from the discovery dataset was frozen and held-out participants were projected onto it using non-negative least squares (NNLS). Explained variance is the proportion of the squared Frobenius norm reconstructed by the factorization. Relative reconstruction error (RRE) = ||V − WH||F / ||V||F. Per-factor Kolmogorov–Smirnov (KS) tests compare the distribution of projected factor loadings in held-out datasets to the discovery dataset; all replication KS *P* > 0.19. Mean loading correlation is the Pearson correlation between mean factor loadings in the training and held-out datasets.

**Supplementary Table 11.** Distribution of NMF archetype assignments.

| Archetype | N (%) |
|---|---|
| Mixed (no dominant factor) | 45,146 (51.8%) |
| F1 (Prudent base) | 25,193 (28.9%) |
| F2 (Cheese & whole grains) | 9,833 (11.3%) |
| F3 (Meat & starchy vegetables) | 1,854 (2.1%) |
| F4 (High sugar & processed foods) | 3,114 (3.6%) |
| F5 (Aromatic vegetables & rice) | 420 (0.5%) |
| F6 (Fruit & low-fat dairy) | 700 (0.8%) |
| F7 (Plant-based & fish) | 902 (1.0%) |

**Supplementary Table 12.** E values for FDR-significant factor X outcome associations.

| Factor | Outcome | HR (95% CI) | *P*-value | FDR *P* | E-value | E-value (CI bound) |
|---|---|---|---|---|---|---|
| F2 (Cheese & whole grains) | CAD | 0.94 (0.90–0.98) | 0.003 | 0.022 | 1.33 | 1.18 |
| F4 (High sugar & processed) | T2D | 1.19 (1.11–1.27) | 9.20e-07 | 7.36e-06 | 1.66 | 1.46 |
| F4 (High sugar & processed) | CKD | 1.15 (1.07–1.23) | 5.15e-05 | 4.12e-04 | 1.56 | 1.35 |
| F7 (Plant-based & fish) | CKD | 0.90 (0.84–0.96) | 8.93e-04 | 0.004 | 1.47 | 1.26 |

E-values quantify the minimum strength of association that an unmeasured confounder would need to have with both the exposure and the outcome to explain away the observed association. E-values and their confidence interval bounds are shown for all factor-outcome associations reaching FDR-corrected significance ($P < 0.05$) in the primary M2 Cox model. HR, hazard ratio; CI, confidence interval; FDR, false discovery rate (Benjamini-Hochberg). E-values were computed using the method of VanderWeele and Ding (2017). An E-value of 1.00 for the CI bound indicates that the confidence interval already includes 1.00.

**Supplementary Table 13.** Clinical biomarker effect sizes in AHEI-matched NMF-discordant pairs (replication dataset).

| Biomarker | Domain | N Pairs | Cohen's d | P | P FDR |
|---|---|---|---|---|---|
| BMI (kg/m²)* | **Body composition** | 594 | 0.215 | <0.001 | 0.006 |
| Body fat % (%)* | | 576 | 0.188 | <0.001 | 0.006 |
| Waist circumference (cm) | | 595 | 0.155 | 0.020 | 0.113 |
| Waist-hip ratio | | 595 | 0.073 | 0.328 | 0.601 |
| Glucose (mmol/L) | **Glycemic** | 437 | -0.009 | 0.710 | 0.923 |
| HbA1c (mmol/mol) | | 516 | -0.039 | 0.831 | 0.925 |
| ApoA (g/L) | **Lipids** | 435 | -0.138 | 0.078 | 0.284 |
| ApoB (g/L) | | 520 | -0.007 | 0.845 | 0.925 |
| HDL-C (mmol/L) | | 441 | -0.173 | 0.029 | 0.138 |
| LDL-C (mmol/L) | | 521 | -0.016 | 0.517 | 0.776 |
| Lipoprotein(a) (nmol/L) | | 339 | -0.115 | 0.172 | 0.406 |
| TG/HDL ratio | | 441 | 0.131 | 0.086 | 0.284 |
| Total cholesterol (mmol/L) | | 524 | -0.054 | 0.308 | 0.597 |
| Triglycerides (mmol/L) | | 523 | 0.092 | 0.114 | 0.342 |
| CRP (mg/L) | **Inflammation** | 525 | 0.156 | 0.015 | 0.098 |
| WBC ($10^9$ cells/L)* | | 560 | 0.260 | <0.001 | 0.001 |
| Albumin (g/L) | **Liver** | 442 | -0.035 | 0.607 | 0.834 |
| ALP (U/L) | | 525 | -0.011 | 0.990 | 0.990 |
| Alpha-1-antitrypsin (g/L) | | 520 | 0.050 | 0.897 | 0.925 |
| ALT (U/L) | | 524 | 0.064 | 0.422 | 0.696 |
| Direct bilirubin (μmol/L) | | 386 | -0.090 | 0.149 | 0.406 |
| GGT (U/L) | | 523 | 0.081 | 0.167 | 0.406 |
| Total bilirubin (μmol/L) | | 517 | -0.097 | 0.081 | 0.284 |
| Creatinine (μmol/L) | **Kidney** | 522 | 0.008 | 0.869 | 0.925 |
| Cystatin C (mg/L) | | 525 | 0.072 | 0.262 | 0.576 |
| Urate (μmol/L) | | 525 | 0.029 | 0.727 | 0.923 |
| Urea (mmol/L) | | 523 | -0.046 | 0.390 | 0.678 |
| Diastolic BP (mmHg) | **Blood pressure** | 582 | 0.040 | 0.503 | 0.776 |
| Systolic BP (mmHg) | | 582 | -0.031 | 0.602 | 0.834 |
| Calcium (mmol/L) | **Other metabolic** | 442 | 0.012 | 0.829 | 0.925 |
| IGF-1 (nmol/L) | | 522 | 0.005 | 0.886 | 0.925 |
| Phosphate (mmol/L) | | 438 | -0.053 | 0.298 | 0.597 |
| Vitamin D (nmol/L)* | | 477 | -0.201 | <0.001 | 0.008 |

Cohen's d represents the standardized mean difference (adverse minus favorable NMF-derived cardiometabolic risk). Positive values indicate higher levels in the adverse group. P values from Wilcoxon signed-rank tests on matched discordant pairs. * Significant after Benjamini-Hochberg FDR correction (P FDR < 0.05).

**Supplementary Table 14.** Clinical biomarker effect sizes in aMED-matched NMF-discordant pairs (replication dataset).

| Biomarker | Domain | N Pairs | Cohen's d | P | P FDR |
|---|---|---|---|---|---|
| BMI (kg/m²)* | **Body composition** | 595 | 0.337 | <0.001 | <0.001 |
| Body fat % (%)* | | 580 | 0.227 | <0.001 | <0.001 |
| Waist circumference (cm)* | | 596 | 0.245 | <0.001 | <0.001 |
| Waist-hip ratio | | 596 | 0.139 | 0.029 | 0.086 |
| Glucose (mmol/L) | **Glycemic** | 460 | -0.011 | 0.660 | 0.751 |
| HbA1c (mmol/mol) | | 512 | -0.014 | 0.922 | 0.922 |
| ApoA (g/L)* | **Lipids** | 455 | -0.192 | 0.004 | 0.015 |
| ApoB (g/L) | | 527 | 0.047 | 0.345 | 0.475 |
| HDL-C (mmol/L)* | | 461 | -0.251 | <0.001 | <0.001 |
| LDL-C (mmol/L) | | 531 | 0.005 | 0.814 | 0.867 |
| Lipoprotein(a) (nmol/L) | | 343 | -0.070 | 0.227 | 0.341 |
| TG/HDL ratio* | | 461 | 0.183 | 0.001 | 0.007 |
| Total cholesterol (mmol/L) | | 530 | -0.057 | 0.512 | 0.630 |
| Triglycerides (mmol/L)* | | 531 | 0.140 | 0.006 | 0.021 |
| CRP (mg/L)* | **Inflammation** | 528 | 0.190 | 0.003 | 0.012 |
| WBC ($10^9$ cells/L)* | | 548 | 0.206 | 0.001 | 0.007 |
| Albumin (g/L) | **Liver** | 462 | -0.183 | 0.021 | 0.068 |
| ALP (U/L) | | 530 | 0.076 | 0.252 | 0.361 |
| Alpha-1-antitrypsin (g/L) | | 528 | -0.019 | 0.417 | 0.551 |
| ALT (U/L) | | 530 | 0.053 | 0.200 | 0.341 |
| Direct bilirubin (μmol/L) | | 393 | 0.128 | 0.188 | 0.341 |
| GGT (U/L) | | 531 | 0.084 | 0.218 | 0.341 |
| Total bilirubin (μmol/L) | | 528 | 0.051 | 0.709 | 0.780 |
| Creatinine (μmol/L) | **Kidney** | 531 | 0.087 | 0.169 | 0.340 |
| Cystatin C (mg/L) | | 530 | 0.123 | 0.175 | 0.340 |
| Urate (μmol/L) | | 527 | 0.130 | 0.065 | 0.153 |
| Urea (mmol/L) | | 530 | -0.071 | 0.213 | 0.341 |
| Diastolic BP (mmHg) | **Blood pressure** | 583 | 0.005 | 0.884 | 0.912 |
| Systolic BP (mmHg) | | 583 | -0.095 | 0.061 | 0.153 |
| Calcium (mmol/L) | **Other metabolic** | 462 | -0.039 | 0.620 | 0.731 |
| IGF-1 (nmol/L) | | 525 | -0.106 | 0.054 | 0.149 |
| Phosphate (mmol/L) | | 460 | -0.044 | 0.516 | 0.630 |
| Vitamin D (nmol/L) | | 473 | -0.109 | 0.102 | 0.224 |

Cohen's d represents the standardized mean difference (adverse minus favorable NMF-derived cardiometabolic risk). Positive values indicate higher levels in the adverse group. P values from Wilcoxon signed-rank tests on matched discordant pairs. * Significant after Benjamini-Hochberg FDR correction (P FDR < 0.05).
